# Genetic variation affects morphological retinal phenotypes extracted from UK Biobank Optical Coherence Tomography images

**DOI:** 10.1101/2020.07.20.20157180

**Authors:** Hannah Currant, Pirro Hysi, Tomas W Fitzgerald, Puya Gharahkhani, Pieter W M Bonnemaijer, UK Biobank Eye and Vision Consortium, International Glaucoma Genetics Consortium, Denize Atan, Tin Aung, Jason Charng, Hélène Choquet, Jamie Craig, Alex W Hewitt, Peng T Khaw, Caroline C W Klaver, Michiaki Kubo, Jue-Sheng Ong, Louis R Pasquale, Charles A Reisman, Mark J Simcoe, Alberta A H J Thiadens, Cornelia M van Duijn, Seyhan Yazar, Eric Jorgenson, Stuart MacGregor, Chris J Hammond, David A Mackey, Janey L Wiggs, Paul J Foster, Praveen J Patel, Ewan Birney, Anthony P Khawaja

## Abstract

Optical Coherence Tomography (OCT) enables non-invasive imaging of the retina and is often used to diagnose and manage multiple ophthalmic diseases including glaucoma. We present the first large-scale quantitative genome-wide association study of inner retinal morphology using phenotypes derived from OCT images of 31,434 UK Biobank participants. We identify 46 loci associated with thickness of the retinal nerve fibre layer or ganglion cell inner plexiform layer. Only one of these loci has previously been associated with glaucoma, and Mendelian randomisation confirms that inner retinal thickness, despite being a valid biomarker for the disease, is not on the same genetic causal pathway as glaucoma. Image analysis methods were used to extract overall retinal thickness at the fovea, representative of hypoplasia, with which three out of the 46 SNPs were associated. These SNPs have been previously linked with pigmentation, confirmed by their association with hair colour in the UK Biobank dataset. We additionally associate these three loci with visual acuity. In contrast to the already known Mendelian causes of severe foveal hypoplasia, our results suggest a previously unknown spectrum of foveal hypoplasia in the population, in part genetically determined, that has consequences on visual function.

## Introduction

The human retina is a highly structured tissue at the back of the eye which converts energy from photons focused by the cornea and lens into neuronal signals to provide vision. The retina is made up of distinct layers of cells, sometimes just one cell thick, which have specific functions in this signal processing. The inner retina, which is closest to the pupil of the eye, is responsible for the final stages of signal transmission through the eye before the signal exits via the optic nerve towards the brain (Figure 1A). Advances in optical coherence tomography (OCT) allow for non-invasive high-resolution imaging of retinal tissue structure, enabling the individual layers to be resolved. The central area of the retina, a region called the macula, has a distinct valley-like morphology that can be clearly seen using OCT. Here, the two inner retinal layers, the retinal nerve fibre layer (RNFL) and ganglion cell inner plexiform layer (GCIPL), taper to non-existence at the centre, the fovea, allowing less light scattering for incoming photons and so providing the region of highest acuity vision (Figure 1B) [1]. The morphology of the macula has been studied previously, with variation in physiology associated with ethnicity [2, 3], age [4, 5] and lifestyle variables such as smoking [6]. There are also several diseases that exhibit changes in the thickness of the inner retina, such as multiple sclerosis [7, 8], diabetic retinopathy [9, 10], Parkinson’s disease [11] and other neurological disorders [12, 13, 14, 15].

**Figure 1.**
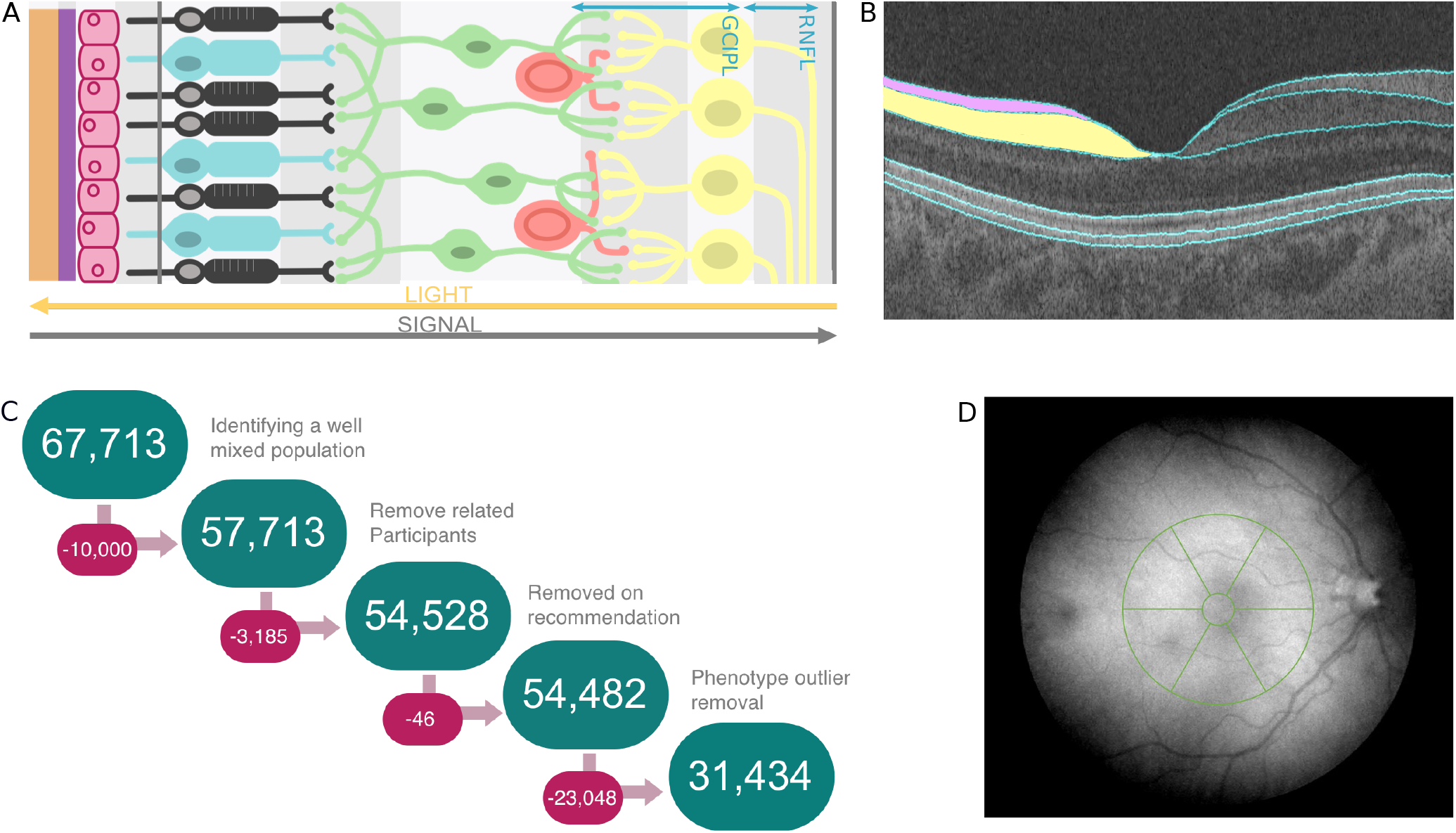
Retinal phenotype data and quality control. A) A diagram illustrating the different retinal layers and the direction of travel for both the light stimulus and the neuronal signal. B) An example of an Optical Coherence Tomography (OCT) image with segmented layer boundaries as labelled by the Topcon Advanced Boundary Segmentation (TABS) algorithm. On the left side of the image, the retinal nerve fibre layer (RNFL) is shaded pink, and the ganglion cell inner plexiform layer (GCIPL) is shaded yellow. C) A schematic of the workflow applied during quality control, involving quality control of both the genotypic and phenotypic data. D) A schematic of the Macula6 grid, a commonly used partition matrix of the macular field when studying the inner retina. The matrix is comprised of 6 sections, with the central field being excluded from analysis.

Most notably, glaucoma, the leading cause of irreversible blindness globally [16], causes thinning of the inner retinal layers [17]. The thickness of the Ganglion Cell Complex (GCC), the collective name for the RNFL and GCIPL, is one of the biomarkers used in diagnosis of primary open angle glaucoma (POAG) [18, 19]. Studies have found genetic variants associated with glaucoma [20, 21, 22], as well as other endophenotypes of the disease such as intraocular pressure (IOP), one of the major risk factors for glaucoma [23, 24]. A number of well-defined loci are known to affect POAG, with over 100 loci reportedly associated with the disease, including at *CDKN2B-AS1, SIX1*/*SIX6, CAV1*/*CAV2, TMCO1* and *GAS7* amongst others [21, 25]. Genetic influence on retinal morphology has been previously studied [26] but largely in smaller cohorts or focused on looking at the retina as a whole [27].

In this study we utilised the large UK Biobank resource where in the latter stages of systematic phenotyping, 67,321 volunteers had OCT imaging centred on the macula [28]. The UK Biobank is a large, well-studied prospective cohort in the UK, with recruitment of adults displaying good general health. On recruitment, participants had a large number of physiological, health and lifestyle-related variables measured. They also consented to on-going linkage of their medical records. The entire 500,000 person cohort has been genotyped and imputed [29]. This dataset therefore provides a well-powered resource to produce a comprehensive view of retinal morphology genetics. In this study we have focused on the morphology of the inner retina, comprising the RNFL and GCIPL, with particular focus on related diseases of these layers, such as POAG. We find 46 loci associated with variation in mean thickness of at least one of these two layers. Many of these loci are associated with other eye traits and broader anthropometric and neurodevelopmental traits. Further interrogation of these SNPs reveals a subset are also associated with foveal hypolasia, the underdevelopment of the foveal dip [30]. The same SNPs are associated with pigmentation and visual acuity. Interestingly, the majority of these loci do *not* coincide with the extensive glaucoma genetic datasets. We show using Mendelian randomisation that IOP, a well-known risk factor for POAG and target of POAG therapy, has strong support for being on the causal pathway of POAG. However genetic variation in inner retinal thickness does not have strong support for a causal relationship with either POAG or IOP. The established use of retinal thickness as a biomarker for POAG is consistent with a change from baseline due to the progression of the disease; however, genetically determined inner retinal thickness largely does not impact development of POAG.

## Results

We performed standard OCT image and genetic quality control to define a high quality and genetically well-mixed subset of the imaged UK Biobank population (see Methods). This resulted in 31,434 people who passed both imaging and genotype filters (Figure 1C). This subset of people had similar sex and age profiles to the overall population (Supplementary Table 2).

We then performed genome-wide association studies (GWAS) of the mean thickness of both the RNFL and GCIPL across the Macula6 grid (Figure 1D). Both GWAS identified many significant loci and showed minimal evidence of inflation (RNFL:*λ* GC = 1.11, Linkage Disequilibrium Score regression (ldsc) Intercept=1.01, Ratio=0.05; GCIPL:*λ* GC = 1.12, ldsc Intercept=1.01, Ratio=0.05, where ratio = (ldsc intercept-1)/(mean(*χ*^2^)-1)), characteristic of a robust GWAS result (Supplementary figure 8). Given that several of the significant loci were common between the two studies, we combined the two GWAS using meta-analysis methods implemented in MTAG [31] (Figure 2). We also explored performing GWAS on the separate thickness measures for each of the Macula6 subfields, or on principle components derived from the Macula6 grid. Due to the large sample size of this study there was little difference in the final list of discovered loci compared to the simpler two-phenotype method. We favoured the simpler two-phenotype model which has fewer parameter choices and is also in-keeping with common clinical usage of these measures.

**Figure 2.**
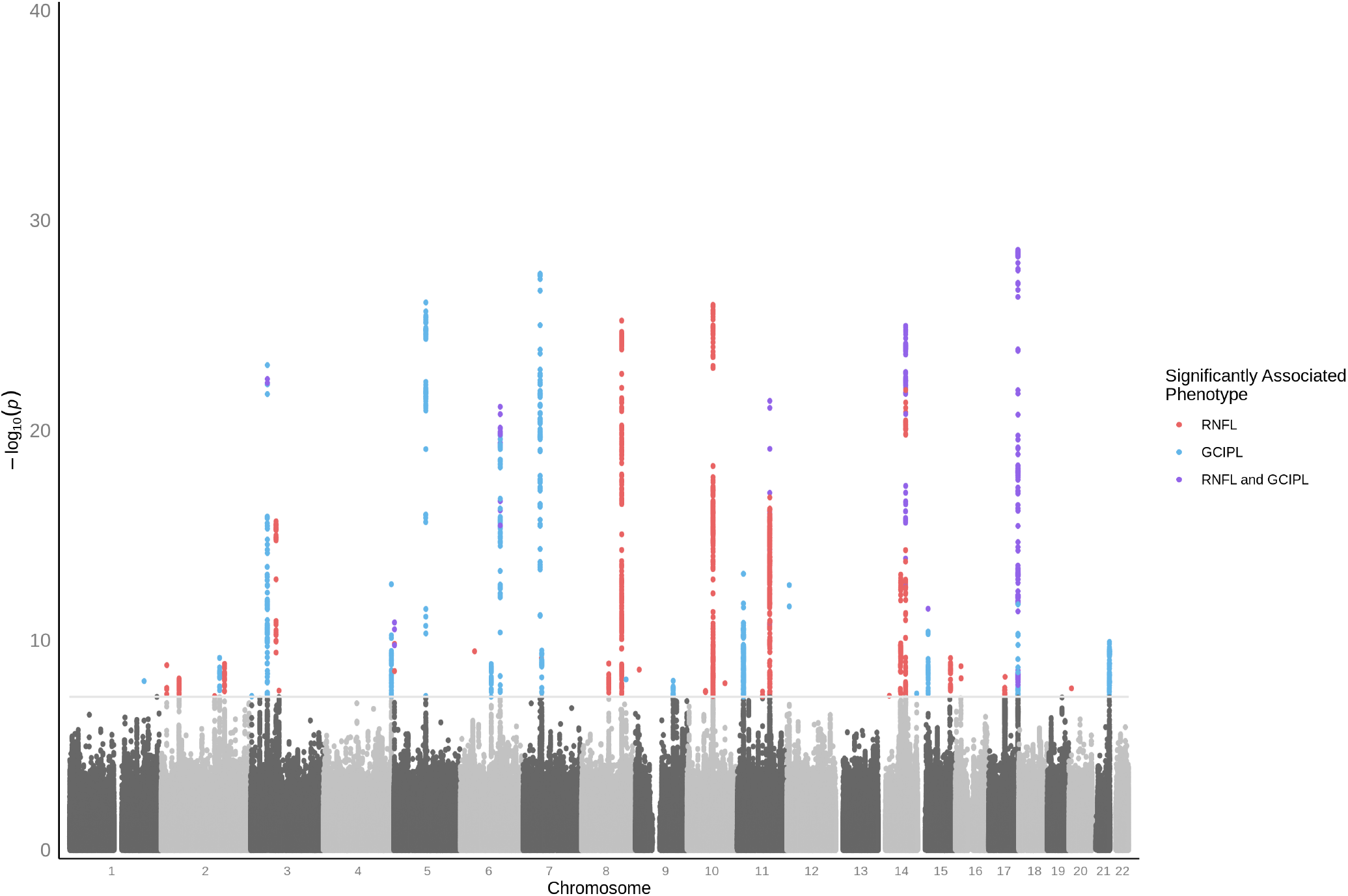
Genome-wide association study of inner retinal thickness phenotypes. Manhattan plot of inner retinal thickness phenotype GWAS p-values, resulting from meta-analysis across RNFL and GCIPL. Variants significantly associated (P <5 × 10^−8^) with only RNFL are highlighted in red, those significantly associated with only GCIPL are highlighted in blue, and those significantly associated with both inner retinal layers are highlighted purple.

There are 46 lead loci discovered across the two phenotypes, listed in Table 1 (additional information available in supplementary Table 3). Many of these loci lie within or near genes which harbour mutations causing a variety of eye phenotypes. These include rs1800407 and rs1042602, at loci previously associated with oculocutaneous albinism (in *OCA2* and *TYR* respectively)[32]. There are also multiple loci that are in or near genes associated with refractive error (*TSPAN10, GNB3, SNAP91, COBL*). Several of the significant loci overlapped with those previously associated with overall retinal thickness [27]. However 36 of our variants are novel and have not previously been associated with retinal morphology. Surprisingly, there was only one locus, rs1254276 at *SIX6*, that was previously associated with POAG [25], though there were several loci that were previously associated with IOP (*TYR, PIK3C2A, NSF, TSPAN10, STOX2*)[24]. Replication was sought in two independent datasets. Replication in the Rotterdam study dataset (Supplementary Table 4) saw correlation of the betas of meta analysed loci associated with RNFL and GCIPL (Pearson’s R = 0.74, P = 1.25 × 10^−6^). In the Raine dataset (Supplementary Table 5), correlation of betas is seen for loci associated with the GCIPL (Pearson’s R = 0.84, P = 5.95 × 10^−6^), however not in RNFL (Pearson’s R = -0.03, P = 0.88). The smaller sample size of both replication data sets meant that many loci would not pass genome-wide significance. The younger age of individuals within the Raine data set may account for the weaker replication. Despite the lack of replication on RNFL in the Raine study, we believe the correlation between GCIPL and RNFL in this study and the association with other retina associated phenotypes indicate that the RNFL results are robust.

**Table 1.**
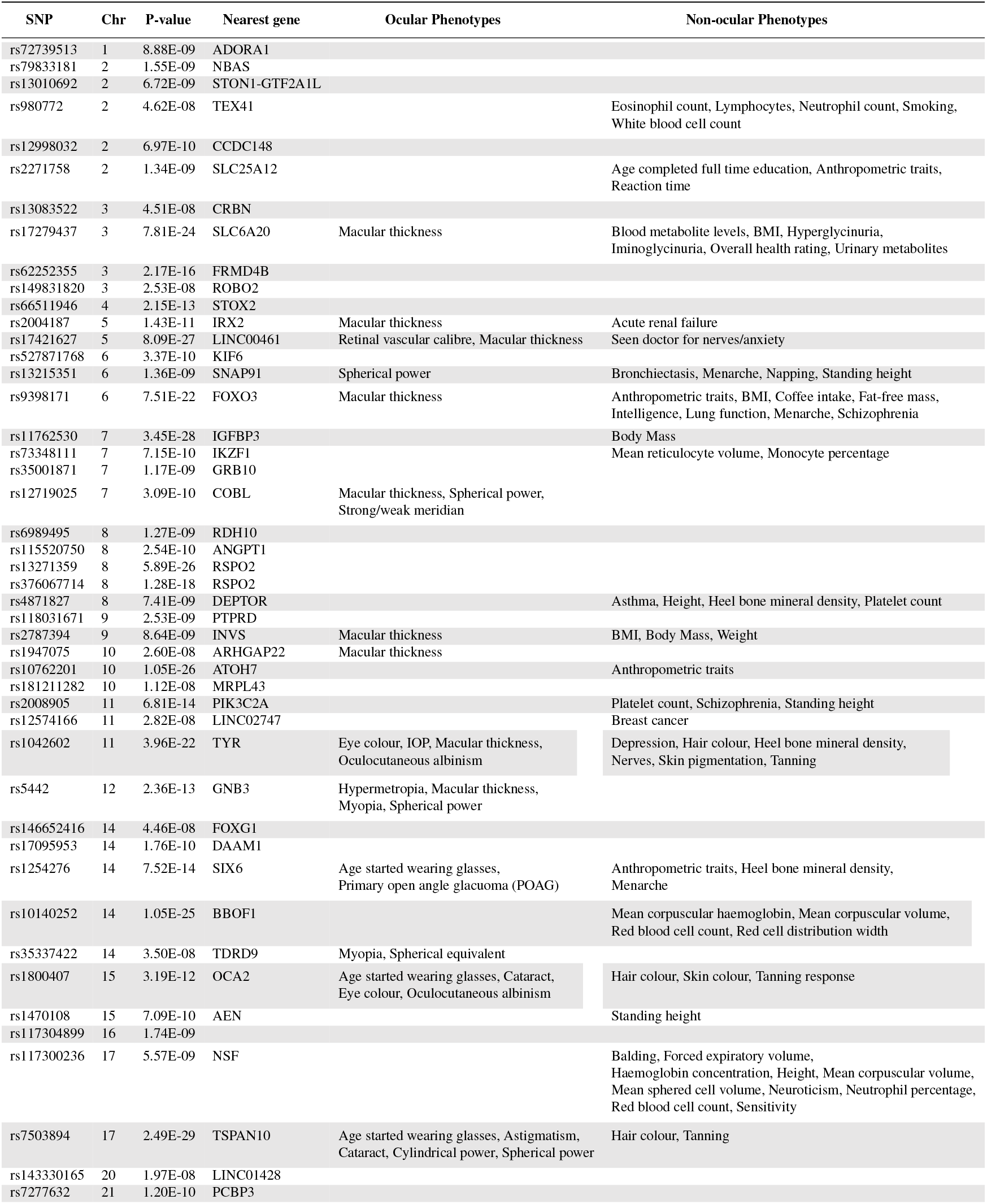
46 SNPs associated with GCIPL or RNFL thickness and annotations of ocular and general biology phenotypes. Variants considered to be representative of a single locus, examples of allelic heterogeneity, are highlighted in the same colour alternating white and grey. For full results including beta values, effect allele specification and standard error please see supplementary Table 3.

**Table 2.** Population Characteristics. Comparison of biological characteristics between whole UK Biobank population with OCT data (n=67,321), the group that passes our quality control criteria (n=31,434), and the group that fails (n=35,887). Results are presented as mean ± standard deviation.

|  | Total Topcon Population | Pass Filter | Fail Filter |
| --- | --- | --- | --- |
| Height (cm) | 168.68 ± 9.25 | 169.43 ± 9.15 | 168.03 ± 9.29 |
| Age (years) | 57 ± 8 | 57 ± 8 | 57 ± 8 |
| Weight (Kg) | 78.12 ± 16.02 | 78.48 ± 15.83 | 77.81 ± 16.18 |
| Sex (m/f) | 29713/34929 | 14837/16597 | 14876/18332 |
| Refractive Error Left (Dioptres) | -0.32 ± 2.73 | 0.23 ± 1.49 | -0.81 ± 3.42 |
| Refractive Error Right (Dioptres) | -0.38 ± 2.73 | 0.19 ± 1.48 | -0.88 ± 3.41 |
| Glaucoma (present/absent) | 6388/60210 (10.61%) | 2751/28314 (9.72%) | 3637/31896 (11.40%) |

**Table 3.** Full GWAS summary statistics for 46 significant variants, including beta, p-value and standard error (SE) for the individual phenotypes, GCIPL and RNFL (labelled accordingly), as well as the values selected within the meta analysis (labelled “MTAG“). A1 is the effect allele.

| SNP | Chr | BP | A1 | A2 | AF | MTAG beta | RNFL beta | GCIPL beta | MTAG p-value | RNFL p-value | GCIPL p-value | MTAG SE | RNFL SE | GCIPL SE |
| --- | --- | --- | --- | --- | --- | --- | --- | --- | --- | --- | --- | --- | --- | --- |
| rs72739513 | 1 | 203080149 | A | G | 0.04 | 0.72 | 0.37 | 0.72 | 8.88E-09 | 1.05E-04 | 8.88E-09 | 0.13 | 0.10 | 0.13 |
| rs12998032 | 2 | 159095496 | C | T | 0.44 | 0.30 | 0.08 | 0.30 | 6.97E-10 | 2.12E-02 | 6.97E-10 | 0.05 | 0.04 | 0.05 |
| rs13010692 | 2 | 48800667 | C | T | 0.32 | 0.22 | 0.22 | 0.09 | 6.72E-09 | 6.72E-09 | 0.09 | 0.04 | 0.04 | 0.05 |
| rs2271758 | 2 | 172701157 | G | T | 0.59 | -0.22 | -0.22 | -0.20 | 1.34E-09 | 1.34E-09 | 3.63E-05 | 0.04 | 0.04 | 0.05 |
| rs79833181 | 2 | 15666802 | C | T | 0.02 | 0.86 | 0.86 | 0.57 | 1.55E-09 | 1.55E-09 | 2.43E-03 | 0.14 | 0.14 | 0.19 |
| rs980772 | 2 | 145442190 | T | G | 0.67 | -0.21 | -0.21 | 0.03 | 4.62E-08 | 4.62E-08 | 0.60 | 0.04 | 0.04 | 0.05 |
| rs13083522 | 3 | 3270368 | G | A | 0.78 | 0.31 | 0.08 | 0.31 | 4.51E-08 | 0.08 | 4.51E-08 | 0.06 | 0.04 | 0.06 |
| rs149831820 | 3 | 77192591 | C | T | 0.06 | -0.42 | -0.42 | -0.31 | 2.53E-08 | 2.53E-08 | 1.48E-03 | 0.07 | 0.07 | 0.10 |
| rs17279437 | 3 | 45814094 | A | G | 0.11 | -0.77 | -0.31 | -0.77 | 7.81E-24 | 1.23E-07 | 7.81E-24 | 0.08 | 0.06 | 0.08 |
| rs62252355 | 3 | 69572006 | C | T | 0.20 | -0.37 | -0.37 | -0.15 | 2.17E-16 | 2.17E-16 | 1.22E-02 | 0.05 | 0.05 | 0.06 |
| rs66511946 | 4 | 184932935 | G | A | 0.40 | -0.36 | -0.11 | -0.36 | 2.15E-13 | 4.36E-03 | 2.15E-13 | 0.05 | 0.04 | 0.05 |
| rs17421627 | 5 | 87847586 | G | T | 0.07 | 0.97 | -0.14 | 0.97 | 8.09E-27 | 4.87E-02 | 8.09E-27 | 0.09 | 0.07 | 0.09 |
| rs2004187 | 5 | 2612747 | C | A | 0.60 | 0.25 | 0.25 | 0.29 | 1.43E-11 | 1.43E-11 | 2.77E-09 | 0.04 | 0.04 | 0.05 |
| rs13215351 | 6 | 84313801 | T | A | 0.25 | -0.33 | -0.06 | -0.33 | 1.36E-09 | 0.12 | 1.36E-09 | 0.05 | 0.04 | 0.05 |
| rs527871768 | 6 | 39419992 | A | G | 0.01 | 1.16 | 1.16 | 0.71 | 3.37E-10 | 3.37E-10 | 3.43E-03 | 0.19 | 0.19 | 0.24 |
| rs9398171 | 6 | 108983527 | T | C | 0.71 | 0.50 | 0.23 | 0.50 | 7.51E-22 | 1.45E-08 | 7.51E-22 | 0.05 | 0.04 | 0.05 |
| rs11762530 | 7 | 46630602 | C | G | 0.59 | 0.53 | 0.07 | 0.53 | 3.45E-28 | 4.77E-02 | 3.45E-28 | 0.05 | 0.04 | 0.05 |
| rs12719025 | 7 | 51100190 | G | A | 0.46 | 0.30 | 0.04 | 0.30 | 3.09E-10 | 0.29 | 3.09E-10 | 0.05 | 0.04 | 0.05 |
| rs35001871 | 7 | 50975439 | C | G | 0.29 | 0.32 | 0.19 | 0.32 | 1.17E-09 | 2.98E-06 | 1.17E-09 | 0.05 | 0.04 | 0.05 |
| rs73348111 | 7 | 50364291 | C | T | 0.01 | 1.12 | 1.12 | 0.66 | 7.15E-10 | 7.15E-10 | 5.70E-03 | 0.18 | 0.18 | 0.24 |
| rs115520750 | 8 | 108739734 | T | G | 0.01 | 1.12 | 1.12 | 0.82 | 2.54E-10 | 2.54E-10 | 4.51E-04 | 0.18 | 0.18 | 0.23 |
| rs13271359 | 8 | 109114426 | T | C | 0.26 | -0.44 | -0.44 | -0.02 | 5.89E-26 | 5.89E-26 | 0.75 | 0.04 | 0.04 | 0.05 |
| rs376067714 | 8 | 109141863 | G | A | 0.18 | -0.48 | -0.48 | -0.11 | 1.28E-18 | 1.28E-18 | 0.13 | 0.05 | 0.05 | 0.07 |
| rs4871827 | 8 | 121061879 | A | G | 0.33 | -0.29 | -0.12 | -0.29 | 7.41E-09 | 2.00E-03 | 7.41E-09 | 0.05 | 0.04 | 0.05 |
| rs6989495 | 8 | 74230223 | T | G | 0.33 | 0.23 | 0.23 | 0.14 | 1.27E-09 | 1.27E-09 | 4.33E-03 | 0.04 | 0.04 | 0.05 |
| rs118031671 | 9 | 10521068 | G | T | 0.01 | 0.93 | 0.93 | 0.76 | 2.53E-09 | 2.53E-09 | 2.21E-04 | 0.16 | 0.16 | 0.21 |
| rs2787394 | 9 | 103007414 | T | C | 0.41 | -0.28 | -0.06 | -0.28 | 8.64E-09 | 0.09 | 8.64E-09 | 0.05 | 0.04 | 0.05 |
| rs10762201 | 10 | 70040111 | G | A | 0.76 | 0.46 | 0.46 | 0.15 | 1.05E-26 | 1.05E-26 | 6.72E-03 | 0.04 | 0.04 | 0.06 |
| rs181211282 | 10 | 102746829 | A | G | 0.03 | 0.67 | 0.67 | 0.24 | 1.12E-08 | 1.12E-08 | 0.11 | 0.12 | 0.12 | 0.15 |
| rs1947075 | 10 | 49741135 | T | C | 0.64 | -0.21 | -0.21 | -0.19 | 2.60E-08 | 2.60E-08 | 1.14E-04 | 0.04 | 0.04 | 0.05 |
| rs1042602 | 11 | 88911696 | A | C | 0.37 | -0.36 | -0.36 | -0.33 | 3.96E-22 | 3.96E-22 | 2.10E-11 | 0.04 | 0.04 | 0.05 |
| rs12574166 | 11 | 69291285 | T | C | 0.15 | 0.28 | 0.28 | 0.24 | 2.82E-08 | 2.82E-08 | 3.92E-04 | 0.05 | 0.05 | 0.07 |
| rs2008905 | 11 | 17184623 | T | C | 0.42 | -0.36 | -0.16 | -0.36 | 6.81E-14 | 1.92E-05 | 6.81E-14 | 0.05 | 0.04 | 0.05 |
| rs5442 | 12 | 6954864 | A | G | 0.07 | -0.69 | -0.08 | -0.69 | 2.36E-13 | 0.27 | 2.36E-13 | 0.09 | 0.07 | 0.09 |
| rs10140252 | 14 | 74528023 | T | G | 0.16 | 0.53 | 0.53 | 0.49 | 1.05E-25 | 1.05E-25 | 1.39E-13 | 0.05 | 0.05 | 0.07 |
| rs1254276 | 14 | 60847001 | T | C | 0.39 | -0.28 | -0.28 | -0.10 | 7.52E-14 | 7.52E-14 | 3.24E-02 | 0.04 | 0.04 | 0.05 |
| rs146652416 | 14 | 29907103 | G | A | 0.03 | 0.62 | 0.62 | 0.45 | 4.46E-08 | 4.46E-08 | 2.44E-03 | 0.11 | 0.11 | 0.15 |
| rs17095953 | 14 | 59719393 | A | G | 0.24 | -0.28 | -0.28 | -0.08 | 1.76E-10 | 1.76E-10 | 0.14 | 0.04 | 0.04 | 0.06 |
| rs35337422 | 14 | 104407243 | C | A | 0.15 | 0.37 | -0.20 | 0.37 | 3.50E-08 | 1.28E-04 | 3.50E-08 | 0.07 | 0.05 | 0.07 |
| rs1470108 | 15 | 89153744 | A | C | 0.34 | 0.24 | 0.24 | 0.21 | 7.09E-10 | 7.09E-10 | 3.77E-05 | 0.04 | 0.04 | 0.05 |
| rs1800407 | 15 | 28230318 | T | C | 0.08 | -0.60 | -0.36 | -0.60 | 3.19E-12 | 4.08E-08 | 3.19E-12 | 0.09 | 0.07 | 0.09 |
| rs117304899 | 16 | 15055042 | G | C | 0.02 | 0.92 | 0.92 | 0.75 | 1.74E-09 | 1.74E-09 | 1.82E-04 | 0.15 | 0.15 | 0.20 |
| rs117300236 | 17 | 44753350 | G | A | 0.72 | -0.25 | -0.25 | -0.30 | 5.57E-09 | 5.57E-09 | 1.24E-07 | 0.04 | 0.04 | 0.06 |
| rs7503894 | 17 | 79583473 | C | T | 0.65 | 0.56 | 0.37 | 0.56 | 2.49E-29 | 3.58E-22 | 2.49E-29 | 0.05 | 0.04 | 0.05 |
| rs143330165 | 20 | 7154672 | T | C | 0.01 | 1.04 | 1.04 | 0.41 | 1.97E-08 | 1.97E-08 | 0.09 | 0.19 | 0.19 | 0.24 |
| rs7277632 | 21 | 47327542 | G | A | 0.72 | 0.34 | 0.09 | 0.34 | 1.20E-10 | 3.56E-02 | 1.20E-10 | 0.05 | 0.04 | 0.05 |

**Table 4.** Association of UK Biobank inner retinal thickness-associated variants with ganglion cell complex thickness in the Rotterdam Study. Comparison between beta and p-values from meta-anlysed inner retinal GWAS (labelled “MTAG“), and GWAS of the GCC thickness in the Rotterdam study (labelled “Rotterdam“).

| SNP | MTAG beta | MTAG p-value | Rotterdam beta | Rotterdam p-value |
| --- | --- | --- | --- | --- |
| rs1042602 | -0.36 | 3.96E-22 | -0.77 | 6.71E-03 |
| rs10762201 | 0.46 | 1.05E-26 | 0.10 | 0.78 |
| rs117300236 | -0.25 | 5.57E-09 | -0.03 | 0.94 |
| rs11762530 | 0.53 | 3.45E-28 | 0.25 | 0.39 |
| rs1254276 | -0.28 | 7.52E-14 | -0.20 | 0.48 |
| rs12574166 | 0.28 | 2.82E-08 | 0.21 | 0.59 |
| rs12719025 | 0.30 | 3.09E-10 | 0.99 | 5.89E-04 |
| rs12998032 | 0.30 | 6.97E-10 | 0.22 | 0.44 |
| rs13010692 | 0.22 | 6.72E-09 | 0.54 | 0.08 |
| rs13083522 | 0.31 | 4.51E-08 | 0.03 | 0.93 |
| rs13215351 | -0.33 | 1.36E-09 | -0.04 | 0.91 |
| rs1470108 | 0.24 | 7.09E-10 | 0.34 | 0.27 |
| rs149831820 | -0.42 | 2.53E-08 | 0.27 | 0.69 |
| rs17279437 | -0.77 | 7.81E-24 | -0.79 | 0.12 |
| rs17421627 | 0.97 | 8.09E-27 | 1.62 | 2.21E-03 |
| rs1800407 | -0.60 | 3.19E-12 | -1.12 | 0.16 |
| rs1947075 | -0.21 | 2.60E-08 | -0.07 | 0.80 |
| rs2004187 | 0.25 | 1.43E-11 | 0.31 | 0.29 |
| rs2008905 | -0.36 | 6.81E-14 | 0.07 | 0.81 |
| rs2271758 | -0.22 | 1.34E-09 | -0.51 | 0.07 |
| rs2787394 | -0.28 | 8.64E-09 | -0.36 | 0.21 |
| rs35001871 | 0.32 | 1.17E-09 | 0.36 | 0.25 |
| rs35337422 | 0.37 | 3.50E-08 | 0.24 | 0.55 |
| rs4871827 | -0.29 | 7.41E-09 | -0.87 | 4.49E-03 |
| rs5442 | -0.69 | 2.36E-13 | 0.38 | 0.46 |
| rs62252355 | -0.37 | 2.17E-16 | -0.27 | 0.43 |
| rs66511946 | -0.36 | 2.15E-13 | -0.51 | 0.11 |
| rs6989495 | 0.23 | 1.27E-09 | 1.00 | 9.50E-04 |
| rs7277632 | 0.34 | 1.20E-10 | 1.35 | 2.25E-05 |
| rs7503894 | 0.56 | 2.49E-29 | 0.57 | 0.06 |
| rs9398171 | 0.50 | 7.51E-22 | 0.69 | 2.55E-02 |
| rs980772 | -0.21 | 4.62E-08 | -0.23 | 0.43 |

**Table 5.** Association of UK Biobank inner retinal thickness-associated variants with retinal nerve fibre layer (RNFL) and ganglion cell inner plexiform layer (GCIPL) thickness in the Raine Study. Comparison between beta and p-values from meta-analysed inner retinal GWAS (labelled “MTAG“), and GWAS of the RNFL and GCIPL in the Raine Study (labelled “Raine RNFL” and “Raine GCIPL“).

| SNP | MTAG beta | MTAG p-value | Raine RNFL beta | Raine RNFL p-value | Raine GCIPL beta | Raine GCIPL p-value |
| --- | --- | --- | --- | --- | --- | --- |
| rs1042602 | -0.36 | 3.96E-22 | -0.20 | 0.11 | - | - |
| rs10762201 | 0.46 | 1.05E-26 | 0.42 | 2.95E-03 | - | - |
| rs115520750 | 1.12 | 2.54E-10 | -0.66 | 0.39 | - | - |
| rs117300236 | -0.25 | 5.57E-09 | -0.15 | 0.28 | - | - |
| rs11762530 | 0.53 | 3.45E-28 | - | - | 0.78 | 8.00E-04 |
| rs118031671 | 0.93 | 2.53E-09 | -0.26 | 0.66 | - | - |
| rs1254276 | -0.28 | 7.52E-14 | -0.24 | 0.05 | - | - |
| rs12574166 | 0.28 | 2.82E-08 | 0.35 | 3.32E-02 | - | - |
| rs12719025 | 0.30 | 3.09E-10 | - | - | 0.57 | 1.66E-02 |
| rs12998032 | 0.30 | 6.97E-10 | - | - | 0.50 | 2.64E-02 |
| rs13010692 | 0.22 | 6.72E-09 | 0.09 | 0.49 | - | - |
| rs13083522 | 0.31 | 4.51E-08 | - | - | -0.28 | 0.32 |
| rs13215351 | -0.33 | 1.36E-09 | - | - | -0.27 | 0.30 |
| rs143330165 | 1.04 | 1.97E-08 | -1.81 | 0.21 | - | - |
| rs146652416 | 0.62 | 4.46E-08 | 0.37 | 0.38 | - | - |
| rs1470108 | 0.24 | 7.09E-10 | 0.10 | 0.43 | - | - |
| rs149831820 | -0.42 | 2.53E-08 | -0.37 | 0.13 | - | - |
| rs17279437 | -0.77 | 7.81E-24 | - | - | -1.76 | 1.53E-06 |
| rs17421627 | 0.97 | 8.09E-27 | - | - | 0.58 | 0.20 |
| rs1800407 | -0.60 | 3.19E-12 | -0.28 | 0.18 | -0.49 | 0.23 |
| rs181211282 | 0.67 | 1.12E-08 | -0.19 | 0.65 | - | - |
| rs1947075 | -0.21 | 2.60E-08 | 0.02 | 0.86 | - | - |
| rs2004187 | 0.25 | 1.43E-11 | 0.20 | 0.10 | 0.45 | 0.06 |
| rs2008905 | -0.36 | 6.81E-14 | - | - | 0.03 | 0.90 |
| rs2271758 | -0.22 | 1.34E-09 | -0.41 | 9.13E-04 | - | - |
| rs2787394 | -0.28 | 8.64E-09 | - | - | -0.69 | 3.71E-03 |
| rs35337422 | 0.37 | 3.50E-08 | - | - | 0.12 | 0.71 |
| rs4871827 | -0.29 | 7.41E-09 | - | - | -0.34 | 0.16 |
| rs5442 | -0.69 | 2.36E-13 | - | - | -1.04 | 2.18E-02 |
| rs62252355 | -0.37 | 2.17E-16 | -0.05 | 0.72 | - | - |
| rs66511946 | -0.36 | 2.15E-13 | - | - | -0.70 | 1.07E-02 |
| rs6989495 | 0.23 | 1.27E-09 | -0.01 | 0.96 | - | - |
| rs72739513 | 0.72 | 8.88E-09 | - | - | 0.26 | 0.71 |
| rs7277632 | 0.34 | 1.20E-10 | - | - | 0.69 | 6.14E-03 |
| rs73348111 | 1.12 | 7.15E-10 | 0.88 | 0.21 | - | - |
| rs7503894 | 0.56 | 2.49E-29 | - | - | 0.38 | 0.14 |
| rs79833181 | 0.86 | 1.55E-09 | -0.06 | 0.92 | - | - |
| rs9398171 | 0.50 | 7.51E-22 | - | - | 0.37 | 0.14 |
| rs980772 | -0.21 | 4.62E-08 | -0.35 | 5.36E-03 | - | - |

**Table 6.** Association of UK Biobank inner retinal thickness-associated variants with primary open-angle glaucoma in the International Glaucoma Genetics Consortium meta-analysis [43]

| SNP | Risk allele | Other allele | Chr | Position | Nearest gene | OR for POAG | P-value |
| --- | --- | --- | --- | --- | --- | --- | --- |
| rs1254276 | T | C | 14 | 60847001 | SIX6 | 1.18 | 2.03E-37 |
| rs117300236 | G | A | 17 | 44753350 | NSF | 0.93 | 1.82E-05 |
| rs5442 | A | G | 12 | 6954864 | GNB3 | 1.11 | 9.97E-05 |
| rs11762530 | C | G | 7 | 46630602 | IGFBP3 | 1.04 | 1.52E-03 |
| rs2787394 | T | C | 9 | 103007414 | INVS | 0.97 | 8.44E-03 |
| rs1042602 | A | C | 11 | 88911696 | TYR | 1.03 | 1.47E-02 |
| rs2008905 | T | C | 11 | 17184623 | PIK3C2A | 0.97 | 2.35E-02 |
| rs7503894 | C | T | 17 | 79583473 | NPLOC4 | 0.97 | 2.56E-02 |
| rs17279437 | A | G | 3 | 45814094 | SLC6A20 | 0.95 | 2.87E-02 |
| rs10762201 | G | A | 10 | 70040111 | ATOH7 | 0.97 | 2.95E-02 |
| rs9398171 | T | C | 6 | 108983527 | FOXO3 | 1.03 | 3.79E-02 |
| rs17421627 | G | T | 5 | 87847586 | LINC00461 | 1.05 | 0.07 |
| rs2004187 | C | A | 5 | 2612747 | IRX2 | 1.02 | 0.14 |
| rs13271359 | T | C | 8 | 109114426 | RSPO2 | 1.03 | 0.18 |
| rs73348111 | C | T | 7 | 50364291 | IKZF1 | 1.08 | 0.19 |
| rs17095953 | A | G | 14 | 59719393 | DAAM1 | 1.03 | 0.24 |
| rs35337422 | C | A | 14 | 104407243 | TDRD9 | 1.02 | 0.25 |
| rs1470108 | A | C | 15 | 89153744 | AEN | 1.02 | 0.27 |
| rs115520750 | T | G | 8 | 108739734 | ANGPT1 | 1.07 | 0.37 |
| rs79833181 | C | T | 2 | 15666802 | NBAS | 1.05 | 0.37 |
| rs62252355 | C | T | 3 | 69572006 | FRMD4B | 1.01 | 0.39 |
| rs12574166 | T | C | 11 | 69291285 | LINC02747 | 0.99 | 0.49 |
| rs13010692 | C | T | 2 | 48800667 | STON1-GTF2A1L | 1.01 | 0.55 |
| rs143330165 | T | C | 20 | 7154672 | LINC01428 | 1.07 | 0.57 |
| rs72739513 | A | G | 1 | 203080149 | ADORA1 | 0.98 | 0.58 |
| rs117304899 | G | C | 16 | 15055042 | NA | 1.05 | 0.62 |
| rs12719025 | G | A | 7 | 51100190 | COBL | 1.01 | 0.63 |
| rs2271758 | G | T | 2 | 172701157 | SLC25A12 | 0.99 | 0.63 |
| rs980772 | T | G | 2 | 145442190 | TEX41 | 0.99 | 0.64 |
| rs181211282 | A | G | 10 | 102746829 | MRPL43 | 0.98 | 0.67 |
| rs4871827 | A | G | 8 | 121061879 | DEPTOR | 1.01 | 0.69 |
| rs13083522 | G | A | 3 | 3270368 | CRBN | 0.99 | 0.69 |
| rs66511946 | G | A | 4 | 184932935 | STOX2 | 1.00 | 0.74 |
| rs118031671 | G | T | 9 | 10521068 | PTPRD | 1.01 | 0.77 |
| rs10140252 | T | G | 14 | 74528023 | BBOF1 | 1.01 | 0.78 |
| rs12998032 | C | T | 2 | 159095496 | CCDC148 | 1.00 | 0.80 |
| rs1800407 | T | C | 15 | 28230318 | OCA2 | 0.99 | 0.81 |
| rs13215351 | T | A | 6 | 84313801 | SNAP91 | 1.00 | 0.86 |
| rs146652416 | G | A | 14 | 29907103 | FOXG1 | 0.99 | 0.86 |
| rs6989495 | T | G | 8 | 74230223 | RDH10 | 1.00 | 0.87 |
| rs7277632 | G | A | 21 | 47327542 | PCBP3 | 1.00 | 0.92 |
| rs1947075 | T | C | 10 | 49741135 | ARHGAP22 | 1.00 | 0.99 |
| rs149831820 | C | T | 3 | 77192591 | ROBO2 | 1.00 | 0.99 |

**Table 7.** Betas and p-values for models of SNP effect on total retinal thickness at the fovea and visual acuity. LogMAR letters beta refers to the difference in number of letters read on a standard LogMAR chart. A1 is the effect allele.

| SNP | A1 | A2 | Foveal thickness<br>A-scan p-value | Foveal thickness<br>B-scan p-value | LogMAR letters<br>p-value | Foveal thickness<br>A-scan beta | Foveal thickness<br>B-scan beta | LogMAR letters<br>beta |
| --- | --- | --- | --- | --- | --- | --- | --- | --- |
| rs7503894 | C | T | 1.26E-23 | 1.00E-25 | 3.54E-19 | -2.35 | -2.43 | -0.30 |
| rs1042602 | A | C | 1.23E-16 | 1.30E-16 | 3.61E-08 | 1.91 | 1.88 | 0.18 |
| rs1800407 | T | C | 3.87E-10 | 2.61E-10 | 2.16E-05 | 2.54 | 2.53 | 0.25 |
| rs17421627 | G | T | 1.36E-02 | 9.22E-03 | 0.06 | 1.05 | 1.09 | 0.07 |
| rs5442 | A | G | 0.07 | 4.35E-02 | 0.81 | 0.80 | 0.88 | 0.01 |
| rs9398171 | T | C | 0.06 | 0.05 | 0.96 | 0.47 | 0.47 | 0 |
| rs12998032 | C | T | 0.06 | 0.06 | 0.66 | 0.42 | 0.41 | 0.02 |
| rs79833181 | C | T | 0.14 | 0.07 | 0.70 | -1.32 | -1.57 | -0.02 |
| rs2008905 | T | C | 0.14 | 0.10 | 0.47 | -0.33 | -0.37 | 0.03 |
| rs13271359 | T | C | 0.14 | 0.11 | 0.23 | 0.38 | 0.40 | -0.05 |
| rs13215351 | T | A | 0.09 | 0.11 | 0.07 | -0.44 | -0.40 | -0.19 |
| rs12719025 | G | A | 0.16 | 0.14 | 0.64 | 0.32 | 0.33 | -0.02 |
| rs6989495 | T | G | 0.40 | 0.15 | 0.58 | -0.20 | -0.33 | 0.02 |
| rs146652416 | G | A | 0.33 | 0.21 | 0.73 | 0.68 | 0.87 | 0.04 |
| rs115520750 | T | G | 0.18 | 0.25 | 0.60 | 1.48 | 1.24 | -0.02 |
| rs1947075 | T | C | 0.40 | 0.27 | 0.75 | -0.20 | -0.25 | -0.04 |
| rs62252355 | C | T | 0.41 | 0.31 | 0.24 | -0.23 | -0.28 | -0.04 |
| rs17279437 | A | G | 0.31 | 0.31 | 0.80 | -0.36 | -0.36 | -0.01 |
| rs117304899 | G | C | 0.32 | 0.32 | 0.75 | -0.94 | -0.93 | 0.01 |
| rs7277632 | G | A | 0.16 | 0.33 | 0.98 | 0.35 | 0.24 | -0.02 |
| rs35337422 | C | A | 0.30 | 0.34 | 0.08 | 0.32 | 0.30 | -0.06 |
| rs181211282 | A | G | 0.42 | 0.38 | 0.07 | 0.58 | 0.63 | 0.08 |
| rs118031671 | G | T | 0.24 | 0.42 | 0.87 | 1.14 | 0.76 | 0 |
| rs72739513 | A | G | 0.64 | 0.44 | 0.58 | 0.28 | 0.45 | 0.08 |
| rs12574166 | T | C | 0.25 | 0.50 | 0.20 | -0.36 | -0.21 | 0.04 |
| rs73348111 | C | T | 0.44 | 0.50 | 0.34 | 0.86 | 0.74 | -0.13 |
| rs35001871 | C | G | 0.47 | 0.51 | 0.53 | 0.18 | 0.16 | -0.05 |
| rs980772 | T | G | 0.57 | 0.53 | 0.63 | -0.14 | -0.15 | -0.01 |
| rs376067714 | G | A | 0.71 | 0.53 | 0.05 | 0.12 | 0.21 | -0.12 |
| rs17095953 | A | G | 0.37 | 0.55 | 3.40E-02 | -0.24 | -0.16 | 0.08 |
| rs66511946 | G | A | 0.68 | 0.58 | 0.57 | -0.10 | -0.13 | -0.02 |
| rs2271758 | G | T | 0.58 | 0.58 | 0.50 | -0.12 | -0.12 | 0.09 |
| rs10140252 | T | G | 0.49 | 0.60 | 0.48 | 0.21 | 0.16 | 0.02 |
| rs4871827 | A | G | 0.67 | 0.64 | 0.67 | -0.10 | -0.11 | -0.03 |
| rs143330165 | T | C | 0.80 | 0.65 | 3.10E-02 | -0.29 | -0.51 | -0.37 |
| rs1470108 | A | C | 0.65 | 0.69 | 0.66 | -0.11 | -0.09 | -0.05 |
| rs2787394 | T | C | 0.77 | 0.70 | 0.09 | 0.06 | 0.08 | -0.25 |
| rs13010692 | C | T | 0.63 | 0.73 | 0.95 | -0.12 | -0.08 | 0 |
| rs10762201 | G | A | 0.59 | 0.75 | 0.14 | -0.14 | -0.08 | 0.09 |
| rs1254276 | T | C | 0.71 | 0.77 | 4.40E-02 | -0.08 | -0.06 | -0.07 |
| rs13083522 | G | A | 0.88 | 0.86 | 0.76 | -0.04 | -0.05 | -0.01 |
| rs11762530 | C | G | 0.97 | 0.89 | 0.40 | -0.01 | -0.03 | 0.04 |
| rs2004187 | C | A | 0.90 | 0.89 | 0.84 | -0.03 | 0.03 | 0 |
| rs149831820 | C | T | 0.95 | 0.93 | 0.17 | 0.03 | -0.04 | -0.06 |
| rs527871768 | A | G | 0.99 | 0.95 | 0.79 | 0.01 | -0.08 | 0.02 |
| rs117300236 | G | A | 0.97 | 0.96 | 1.10E-02 | 0.01 | 0.01 | -0.10 |

**Table 8.**
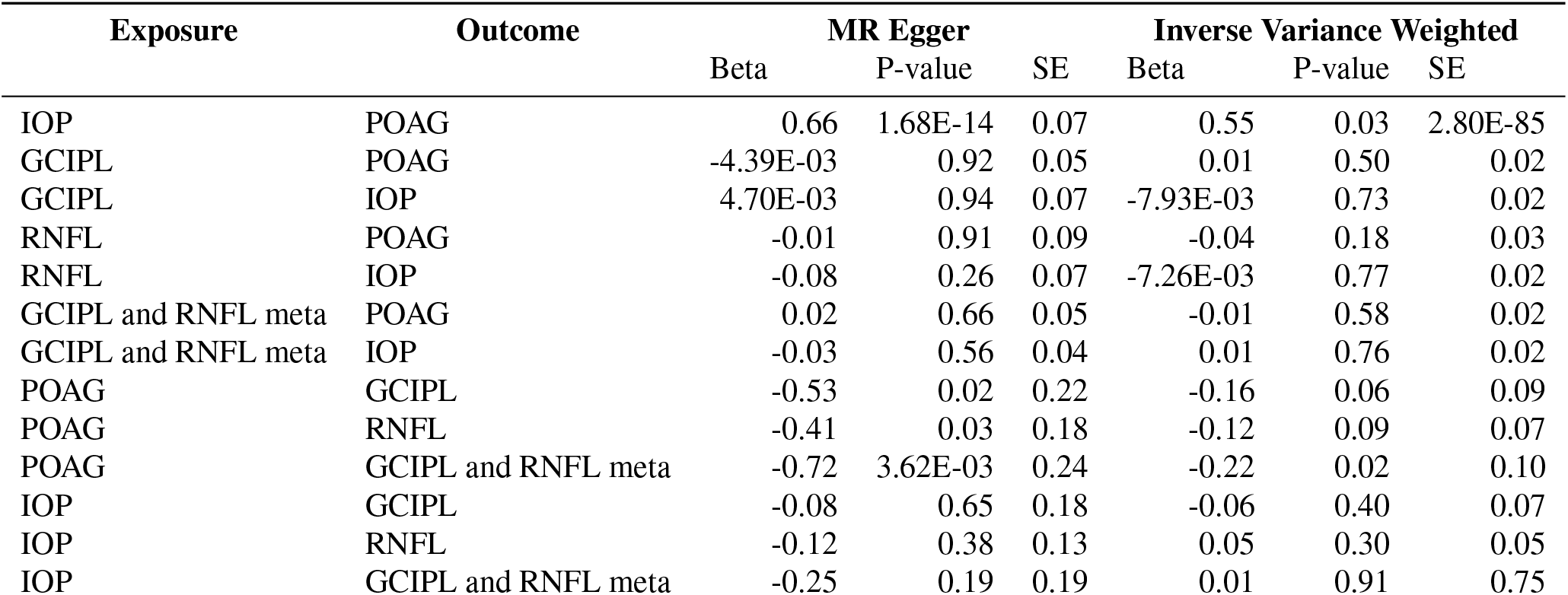
Results of bidirectional two-sample Mendelian randomisation analysis between IOP and POAG, IOP and retinal layer thickness, and POAG and retinal layer thickness. Values reported for two meta analysis methods, MR Egger and Inverse Variance Weighted. POAG summary statistics were taken from the POAG International Glaucoma Genetics Consortium (IGGC) meta analysis [43]. Summary statistics for genetic association studies of IOP, were taken from [24].

| Exposure | Outcome | MR Egger |  |  | Inverse Variance Weighted |  |  |
| --- | --- | --- | --- | --- | --- | --- | --- |
|  |  | Beta | P-value | SE | Beta | P-value | SE |
| IOP | POAG | 0.66 | 1.68E-14 | 0.07 | 0.55 | 0.03 | 2.80E-85 |
| GCIPL | POAG | -4.39E-03 | 0.92 | 0.05 | 0.01 | 0.50 | 0.02 |
| GCIPL | IOP | 4.70E-03 | 0.94 | 0.07 | -7.93E-03 | 0.73 | 0.02 |
| RNFL | POAG | -0.01 | 0.91 | 0.09 | -0.04 | 0.18 | 0.03 |
| RNFL | IOP | -0.08 | 0.26 | 0.07 | -7.26E-03 | 0.77 | 0.02 |
| GCIPL and RNFL meta | POAG | 0.02 | 0.66 | 0.05 | -0.01 | 0.58 | 0.02 |
| GCIPL and RNFL meta | IOP | -0.03 | 0.56 | 0.04 | 0.01 | 0.76 | 0.02 |
| POAG | GCIPL | -0.53 | 0.02 | 0.22 | -0.16 | 0.06 | 0.09 |
| POAG | RNFL | -0.41 | 0.03 | 0.18 | -0.12 | 0.09 | 0.07 |
| POAG | GCIPL and RNFL meta | -0.72 | 3.62E-03 | 0.24 | -0.22 | 0.02 | 0.10 |
| IOP | GCIPL | -0.08 | 0.65 | 0.18 | -0.06 | 0.40 | 0.07 |
| IOP | RNFL | -0.12 | 0.38 | 0.13 | 0.05 | 0.30 | 0.05 |
| IOP | GCIPL and RNFL meta | -0.25 | 0.19 | 0.19 | 0.01 | 0.91 | 0.75 |

In addition to the *TYR, OCA2* and *TSPAN10* loci, which we discuss in more depth below, many of the loci associated in this study are close to well established genes involved in other aspects of ocular biology. Associated loci include rs10762201 and rs2004187 near the *ATOH7* and *IRX2* genes respectively, both associated with eye development [33, 34], rs79833181 near *NBAS* associated with optic atrophy [35], rs149831820 near *ROBO2* associated with retinal ganglion cell axon guidance [36, 37], and rs73348111 (at *IKZF1*), rs13271359 and rs376067714 (both at *RSPO2*) involved in differentiation and retinal cell definition [38, 39]. These SNPs and the others in Table 1 show the link between common variation and the large effects of either Mendelian disease or animal models in eye biology, and provide both feasible paths for more exploration of the biology and the impact of natural population variation on specific aspects of eye biology.

Several of the loci we identified were previously associated with refractive error, despite us adjusting for refractive error in our statistical models. However, the two most established loci associated with refractive error, at *LAMA2* (rs12193446) and *GJD2* (rs524952) [40], were not significantly associated with inner retinal morphology in our MTAG analysis (P=0.32 and P=0.12, respectively). This suggests that there are some shared genetic processes between myopia and inner retinal morphology and that these results are not being driven by residual confounding or magnification artefact due to refractive error.

The presence of loci associated with occulocutaneous albinism, whose effect on foveal morphology has been well documented [41], prompted us to examine an additional measure of retinal morphology. We created a simple model of the total retinal thickness across the macula from the raw OCT images, registered to the foveola (see Methods), and saw how such models varied when stratified by the genotype at each of the 46 lead loci. This revealed that some variants have a more notable diffuse effect across the whole scanned retinal area, while others have a predominant effect on the fovea (Figure 3 and Supplementary Figure 9). To produce a statistical value for this difference, a linear model was constructed modelling the effect of each of the discovery set loci on the total thickness of the retina at the foveola. Three SNPs showed significant differences in retinal thickness at the foveola: rs7503894 (*TSPAN10*, P=1.00 × 10^−25^), rs1042602 (*TYR*, P=1.23 × 10^−16^) and rs1800407 (*OCA2*, P=2.61 × 10^−10^).

**Figure 3.**
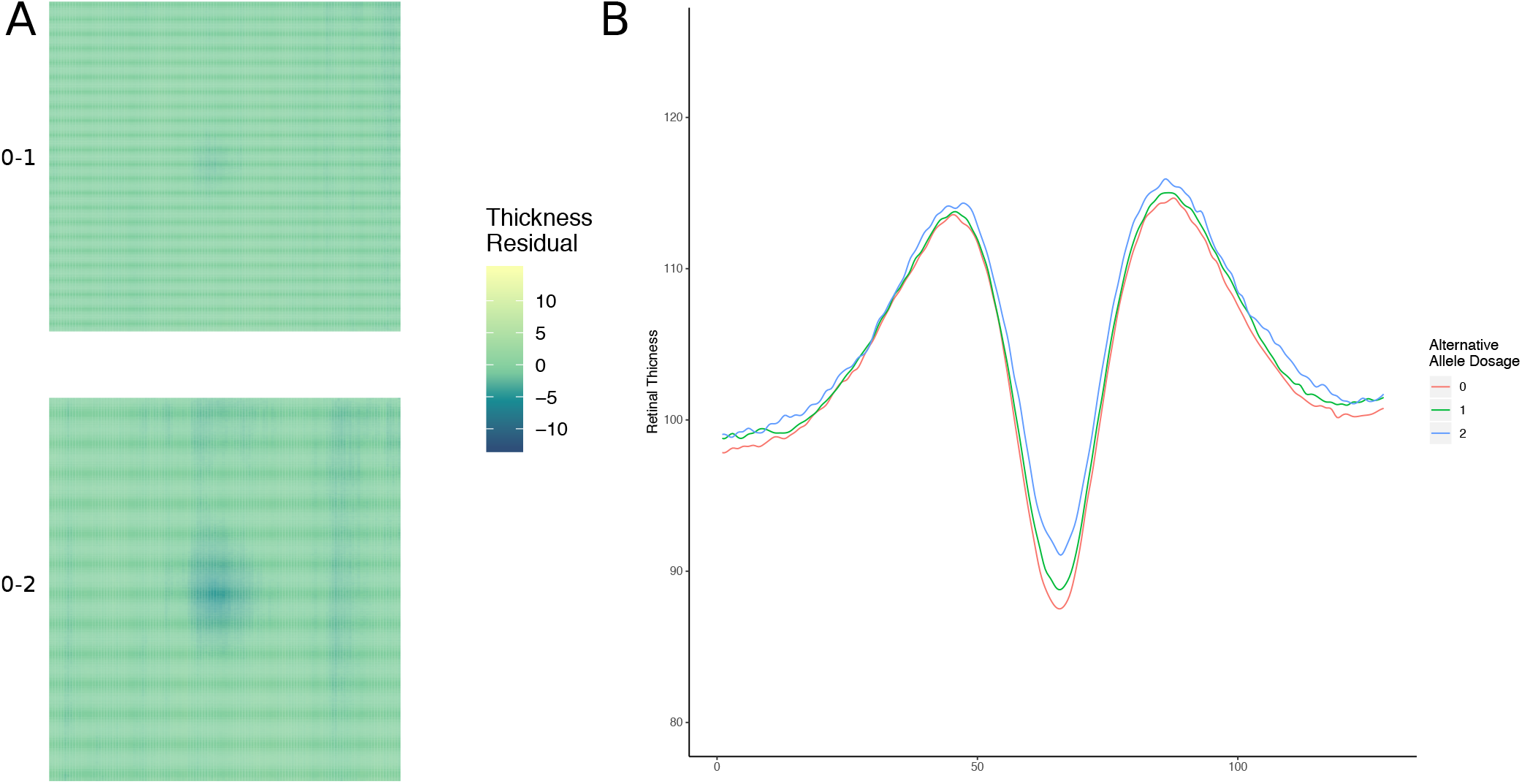
Higher dimensional detailed retinal phenotyping. A) A model of the macular field showing the difference in mean total retinal thickness between those with homozygous reference and heterozygous alleles (top), and homozygous reference and homozygous alternative alleles (bottom) at rs1042602 (*TYR*). B) Models of the mean overall retinal thickness across three groups defined by their allele state, homozygous reference (0), heterozygous (1) or homozygous alternative (2) at rs1042602 (*TYR*). The y axis represents total retinal thickness.

Due to the pre-established link of these SNPs with pigmentation, the effect of the three foveal thickness-associated SNPs on both hair colour and skin colour was explored using a linear model (Figure 4). All three loci were significantly associated with hair colour (*TSPAN10*: P=1.27 × 10^−9^, *TYR*: P=5.08 × 10^−12^, *OCA2*: P=4.82 × 10^−23^). The direction of effect of hair colour change (ranking hair colour from light to dark; see Methods) relative to the direction of effect on foveal hypoplasia was inconsistent across the loci. For the *TYR* and *TSPAN10* loci, the allele associated with greater foveal hypoplasia was also associated with lighter hair colour. However, for the *OCA2* locus, the allele associated with greater foveal hypoplasia was associated with darker hair colour. Only the well-described oculocutaneous albinism variants, *TYR* and *OCA2*, were significantly associated with skin colour (P=2.19 × 10^−4^ & 2.31 × 10^−3^, respectively). There are only 19 individuals in UK Biobank with documented albinism from the Hospital Episode Statistics (ICD10 codes); this is likely an underestimate due to under-reporting and diagnosis, but the frequency of the risk variants at these loci are far higher than the documented levels of oculocutaneous albinism in the UK population, suggesting that the majority of these people are not clinically classified as having eye defects due to the condition. These results show that common natural variation in pigmentation pathways influence retinal development across broad populations but with complex, pleiotropic effects (see Discussion).

**Figure 4.**
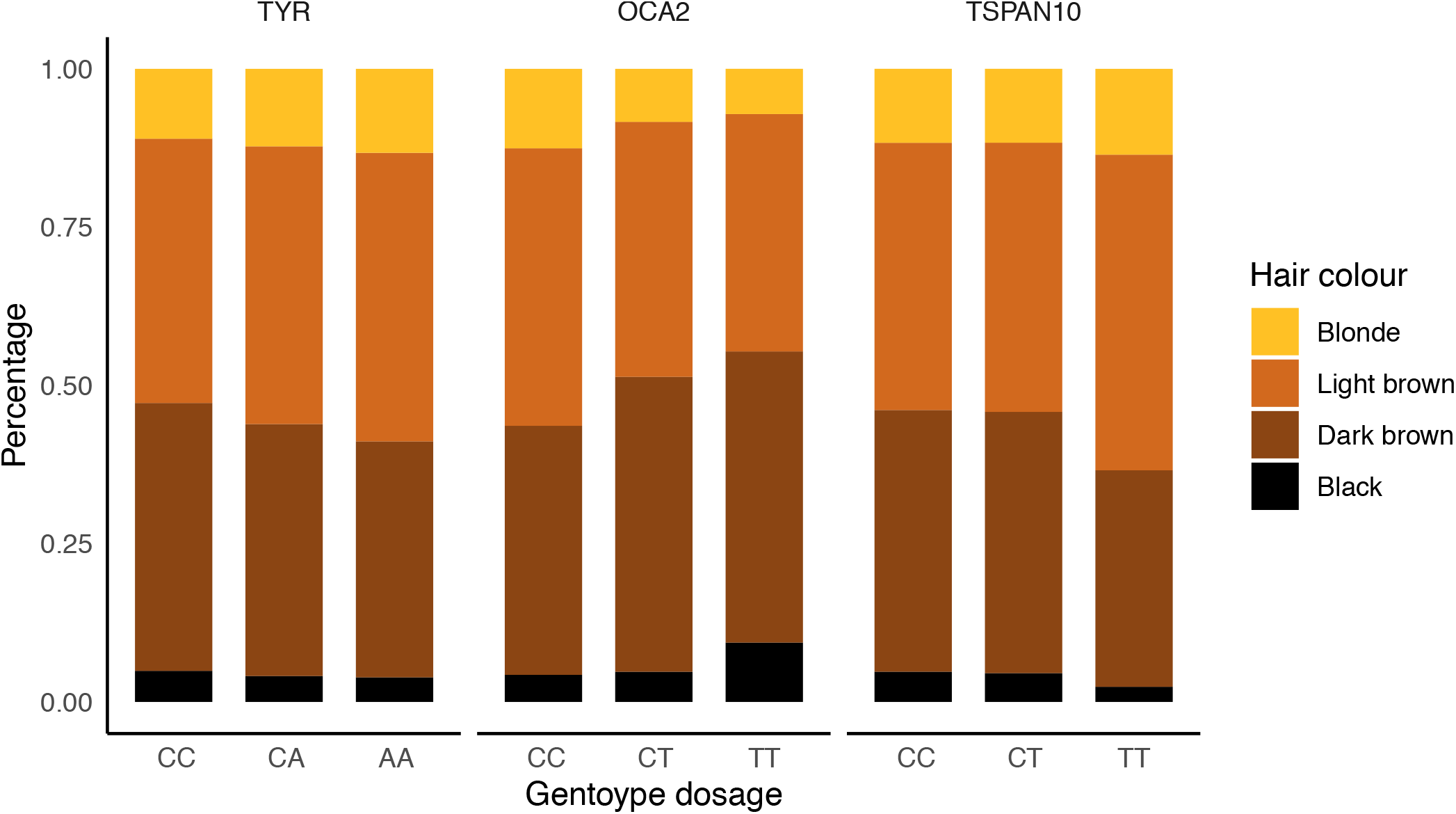
Hair colour stratified by genotype state. Proportions of self-reported hair colour within our dataset population plotted stratified by genotype at the three overall retinal thickness associated loci. Genotypes are aligned so the allele on the right cause a thicker foveola. From left to right: *TYR* (rs1042602), *OCA2* (rs1800407), *TSPAN10* (rs7503894).

To determine whether any of the retinal morphology-associated variants additionally had effects on retinal function, we examined their association with visual acuity (Supplementary Table 7). Three of the 46 variants were significantly associated with visual acuity at a Bonferroni-corrected threshold (P = 3.54 × 10^−19^, 3.61 × 10^−8^ and 2.16 × 10^−5^ for variants at *TSPAN10, TYR* and *OCA2* respectively). Interestingly, these 3 loci are also the loci associated with foveal hypoplasia. The relationship between foveal hypoplasia and visual acuity had a consistent direction across the three variants; the allele associated with a greater degree of foveal hypoplasia was associated with worse visual acuity.

To further explore the underlying biological mechanisms and pathways associated with the traits, we used GARFIELD [42] that associates the full spectrum of GWAS loci with regulatory features from different cell or tissue types. There is an over 10-fold enrichment for loci associated with either RNFL or GCIPL (P <0.05) in eye tissues (Figure 5). Similar-fold enrichment is seen in pancreas (>15x), kidney (>10x), blood (>15x) and brain (>15x) (See Discussion).

**Figure 5.**
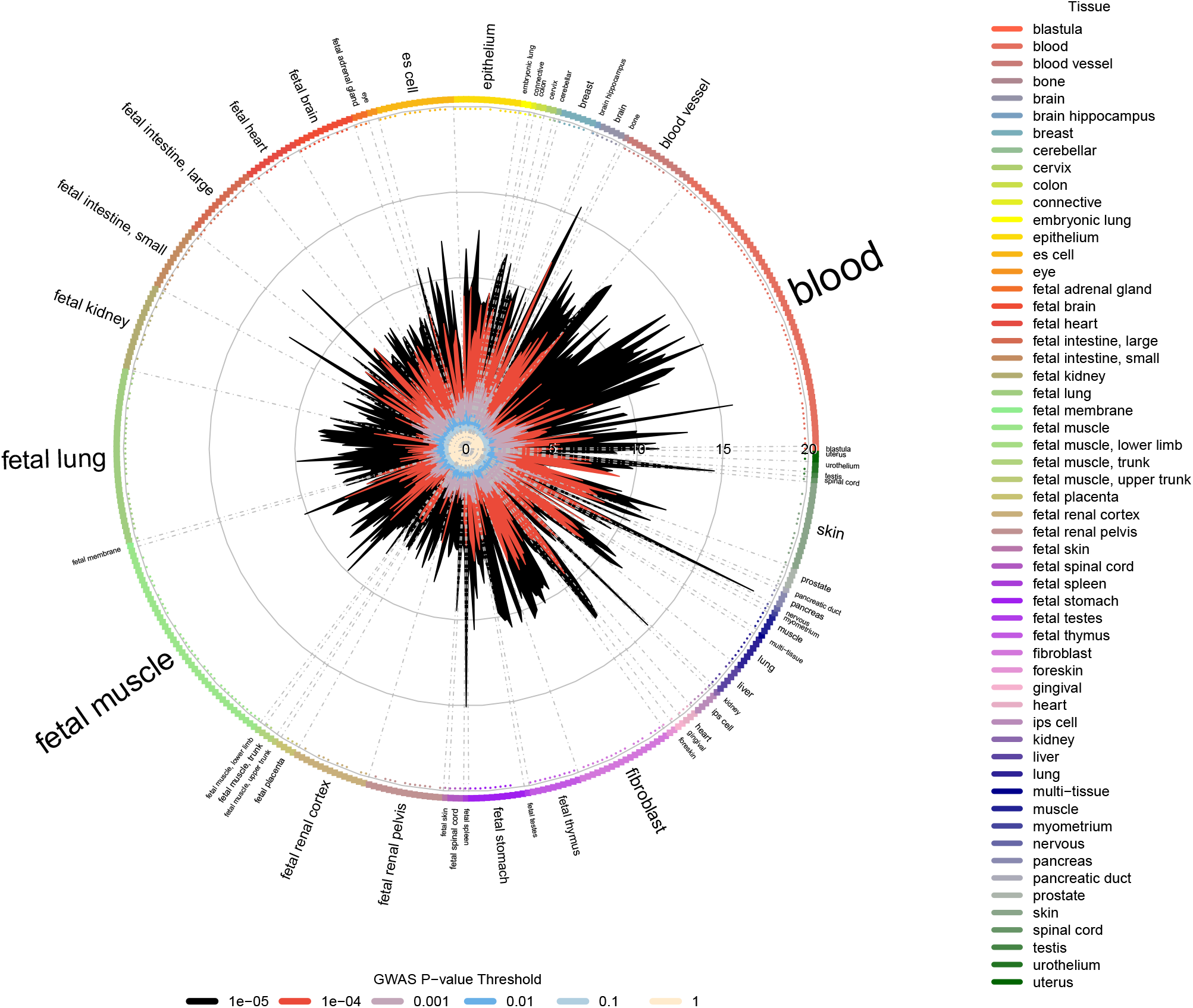
Regulatory feature association using GARFIELD. Wheel plot of enrichment analysis on meta analysed GCIPL and RNFL GWAS results across a number of cell types, as performed in GARFIELD. Associations at different GWAS P-value thresholds are represented in different colours

Prompted by the surprising lack of overlap between the inner retinal phenotypes and the established inner retinal disease of glaucoma, we performed two-sample Mendelian randomisation studies between the retinal morphology traits (RNFL and GCIPL), IOP and POAG. Mendelian randomisation is a statistical technique that uses genetics to test a suspected causal relationship between an “exposure” variable (in this case IOP, RNFL or GCIPL thickness) and an outcome variable (in this case POAG, with summary statistics taken from the International Glaucoma Genetics Consortium (IGGC) meta-analysis [43], or IOP, with summary statistics taken from [24]). As expected, there was evidence for a strong causal link between IOP and POAG, with strong concordance of the effects of variants on IOP with the effects on POAG (Figure 6A & B). This concordance is consistent with evidence that lowering IOP reduces the risk of the progression of POAG [44]. In contrast, there was no evidence of a causal effect of either GCIPL or RNFL on IOP or POAG (Figure 6C & D, Supplementary figures 10 & 11). Conducting the analysis with POAG or IOP as the exposure, and retinal thickness as the outcome, there was no evidence for an effect of IOP on retinal layer thickness, and at most weak support via one meta-analysis technique for a causal relationship between POAG and retinal thickness of both GCIPL and RNFL (Supplementary Figure 12 & 13, Supplementary Table 8); given the strong causal link of IOP to POAG shown by both MR and drug treatments, if this causal link between POAG and inner retinal thickness is present, this data indicates that it is not on the causal pathway with IOP, and likely takes a different biological route.

**Figure 6.**
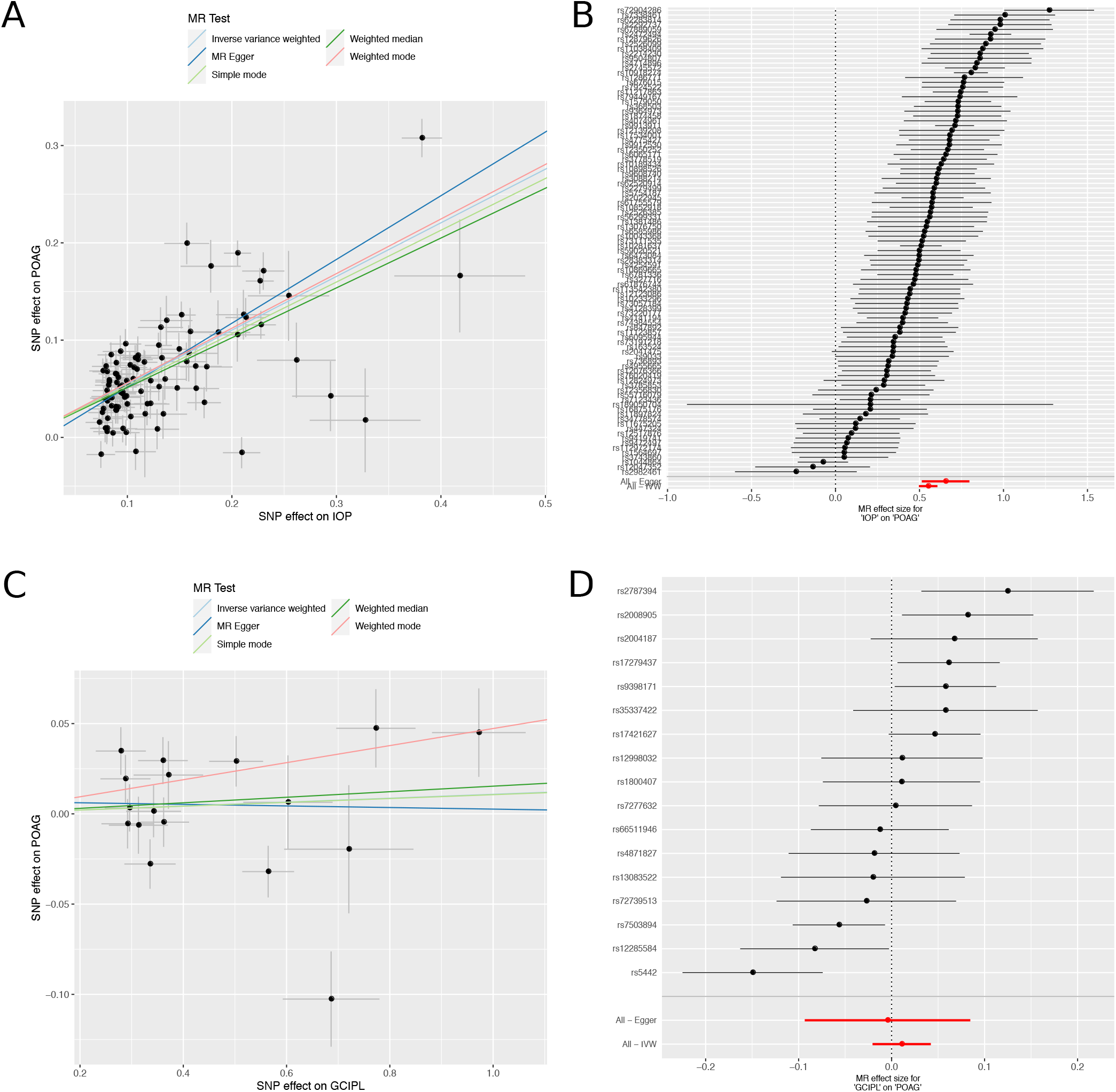
Mendelian Randomisation Analysis. A) Scatter plot of the relationship between the effect size of SNPs found significantly associated to intraocular pressure (IOP), and the effect of those SNPs on primary open angle glaucoma (POAG). B) Forest plot showing effect size and direction of SNPs significantly associated with IOP on POAG. C) Scatter plot of the relationship between the effect size of SNPs found significantly associated with ganglion cell inner plexiform layer (GCIPL) thickness, and the effect of those SNPs on POAG. D) Forest plot showing effect size and direction of SNPs significantly associated with the thickness of the GCIPL on POAG. POAG summary statistics were taken from the POAG International Glaucoma Genetics Consortium (IGGC) meta analysis [43]. Summary statistics for genetic association studies of IOP, were taken from [24].

**Figure 7.**
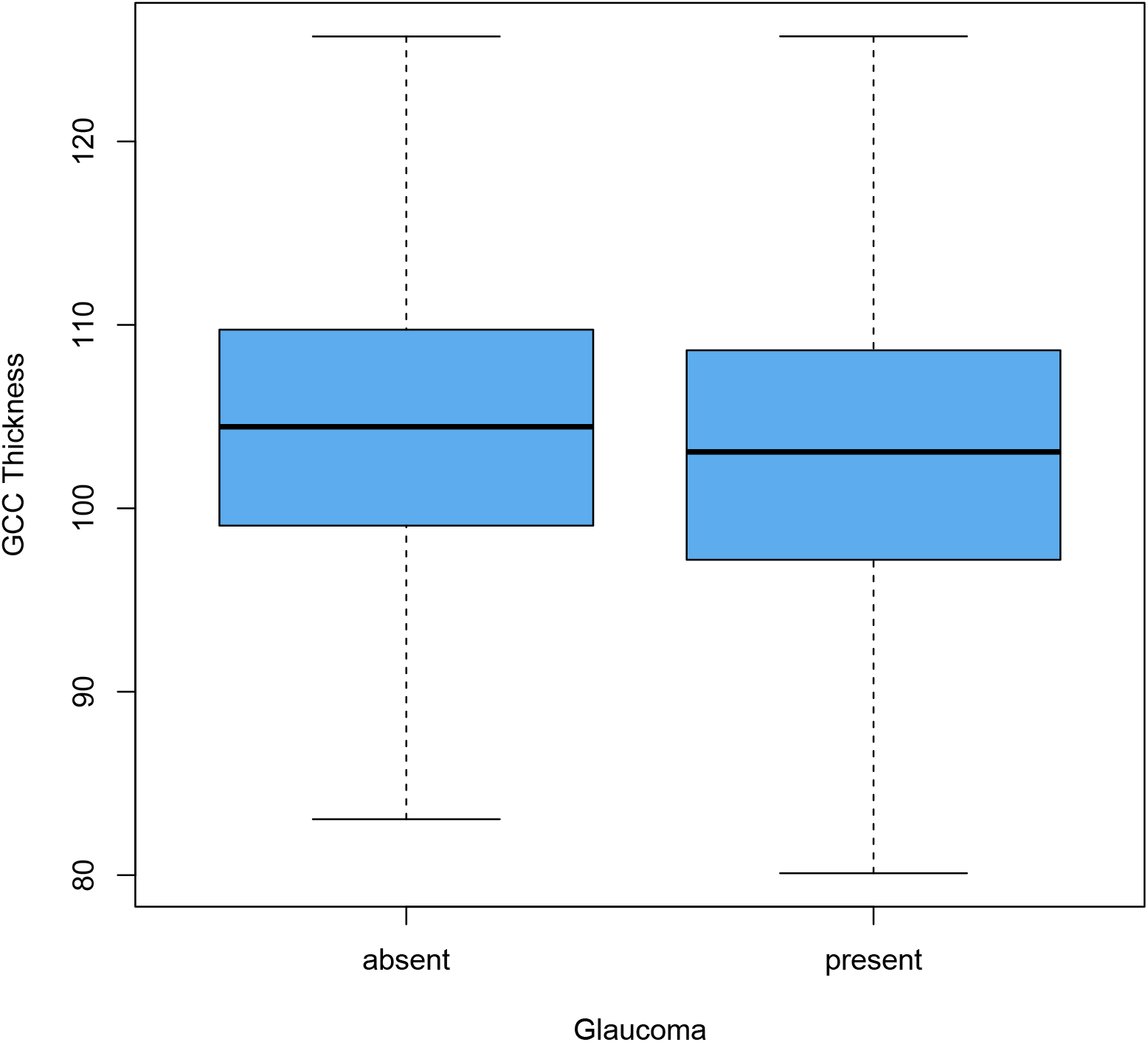
GCC thickness in the UK Biobank glaucoma population. A boxplot showing the thickness of the ganglion cell complex in the dataset comparing those who have glaucoma (n=2751), to those that do not (n=28,314). An accompanying Wilcoxon rank sum test was performed that showed a statistically significant difference between the two populations (P = 2.2 × 10^−16^).

**Figure 8.**
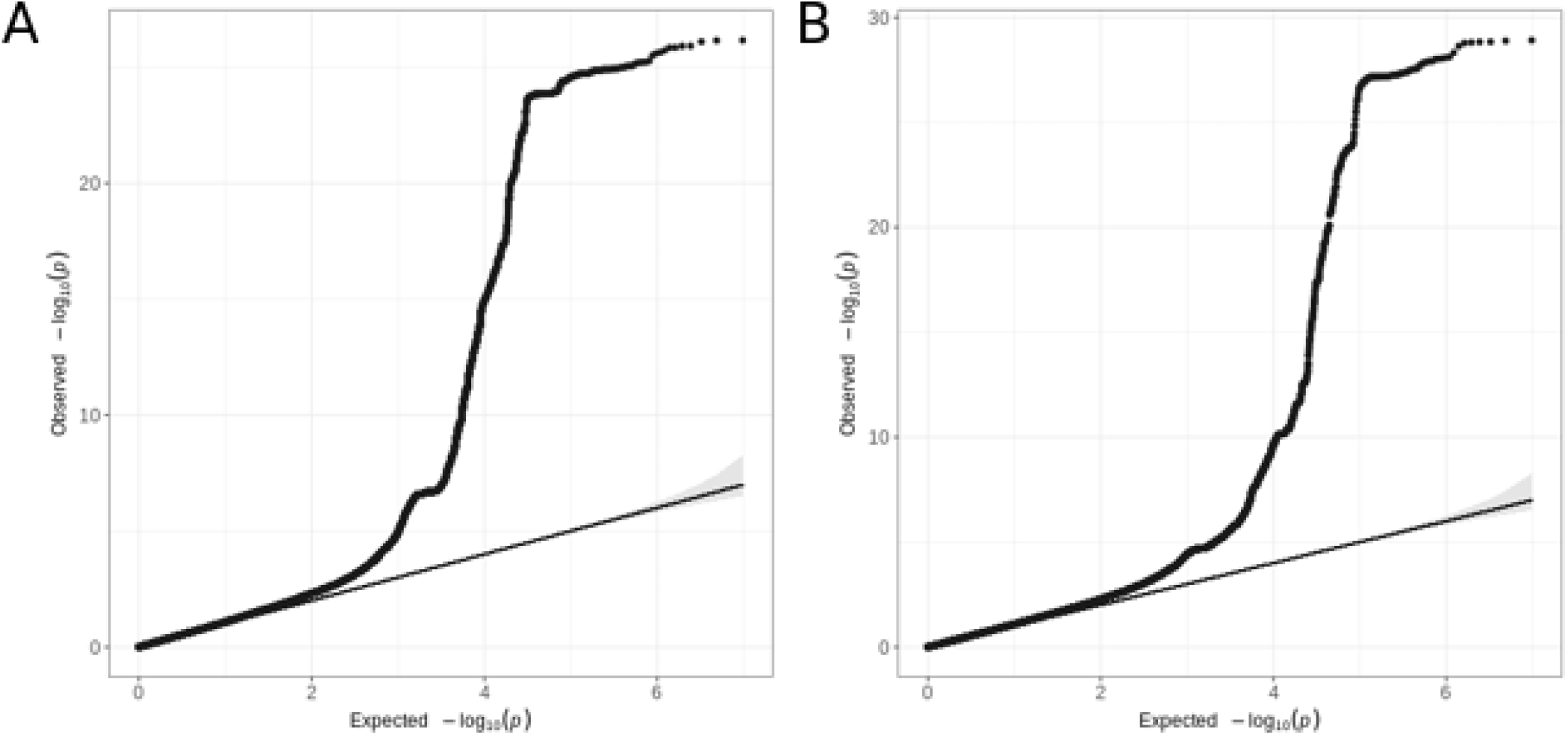
Quantile-quantile plots of retinal thickness GWAS. A) The quantile-quantile plot (qq-plot) for the GWAS of RNFL thickness prior to meta analysis (Lambda GC = 1.11, Intercept = 1.01, Ratio = 0.05). B) The qq-plot for the GWAS of GCIPL thickness prior to meta analysis (Lambda GC = 1.12, Intercept = 1.01, Ratio = 0.05).

**Figure 9.**
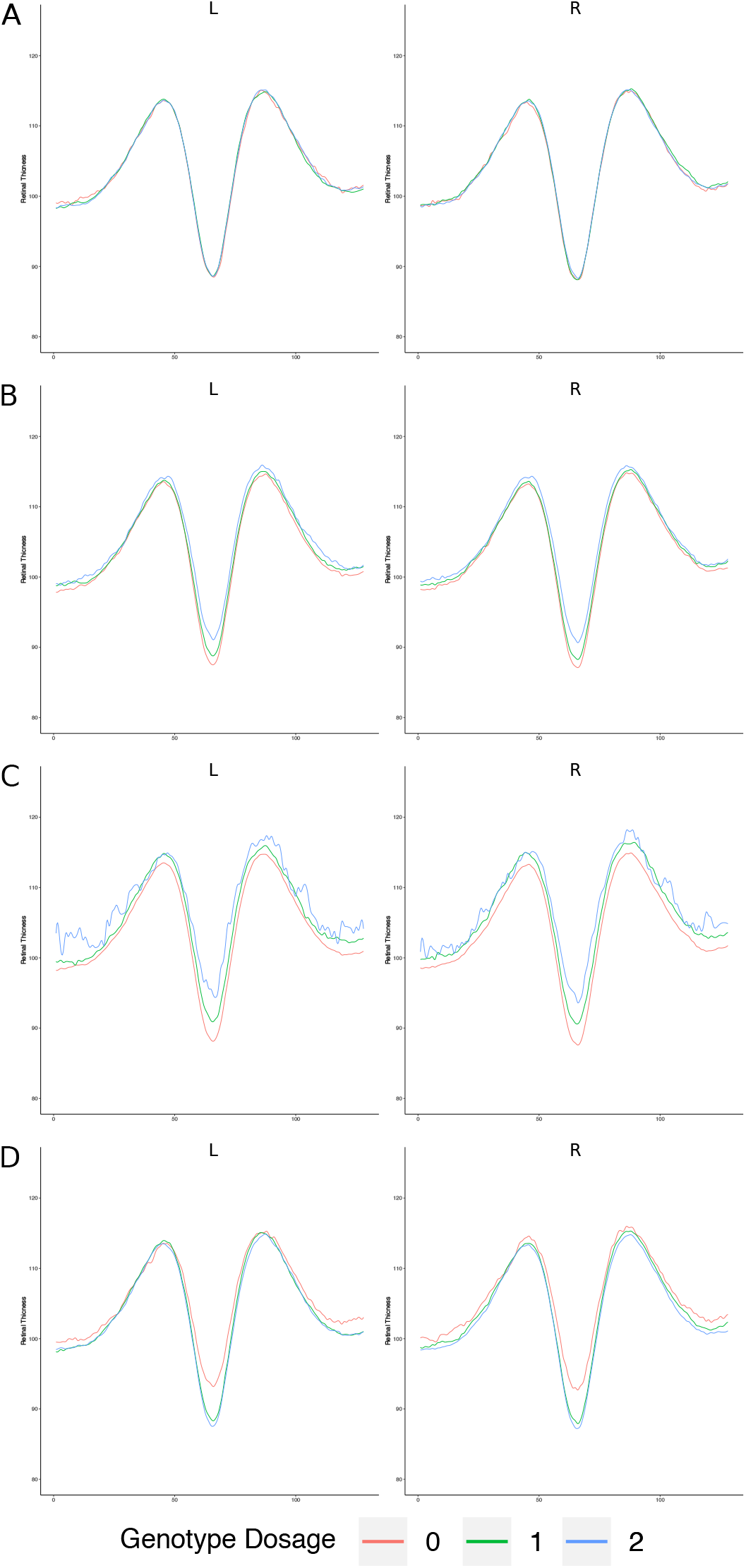
Higher dimensional detailed retinal phenotyping. Mean cross-sectional models of overall retinal thickness across three groups defined by their allele state, homozygous reference, heterozygous or homozygous alternative at: A) *IGFBP3* (rs11762530), the variant with the lowest P-value aside from *TSPAN10*, as a control B) *TYR* (rs1042602) C) *OCA2* (rs1800407) D) *TSPAN10* (rs7503894). The y axis is representative of overall retinal thickness.

**Figure 10.**
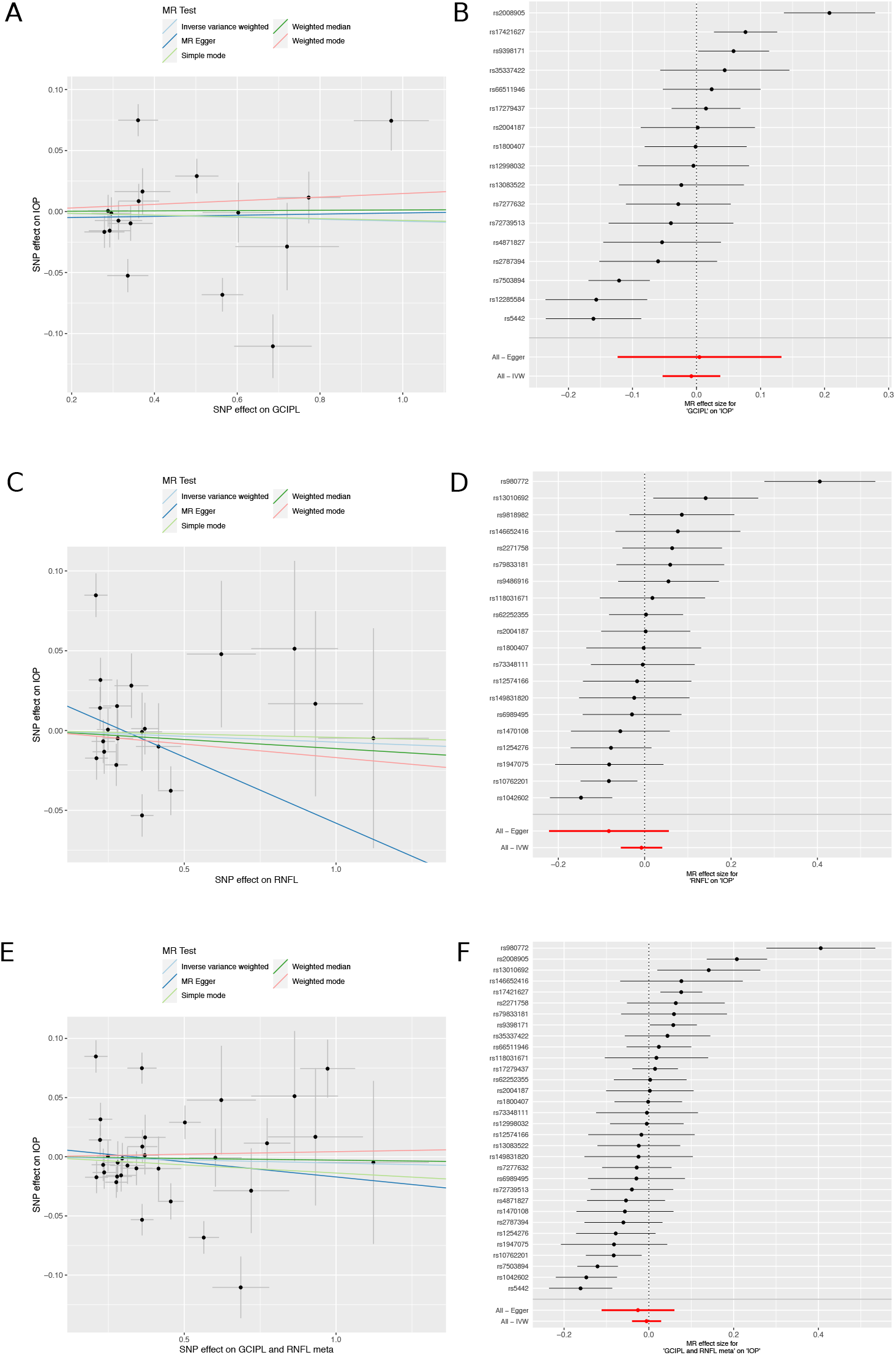
Mendelian Randomisation of Retinal Layers and intraocular pressure. A) Scatter plot of the relationship between the effect size of SNPs found significantly associated with GCIPL thickness, and the effect size of those SNPs on IOP. B) Forest plot showing the effect size and direction of SNPs significantly associated with GCIPL thickness on IOP. C) Scatter plot of the relationship between the effect size of SNPs found significantly associated with RNFL thickness, and the effect of those SNPs on IOP. D) Forest plot showing the effect size and direction of SNPs significantly associated with RNFL thickness on IOP. E) Scatter plot of the relationship between the effect size of SNPs found significantly associated in the meta analysed GCIPL and RNFL thicknesses, and the effect of those SNPs on IOP. F) Forest plot showing the effect size and direction of SNPs significantly associated in the meta analysed GCIPL and RNFL thickness on IOP. Summary statistics for genetic association studies of IOP, were taken from [24].

**Figure 11.**
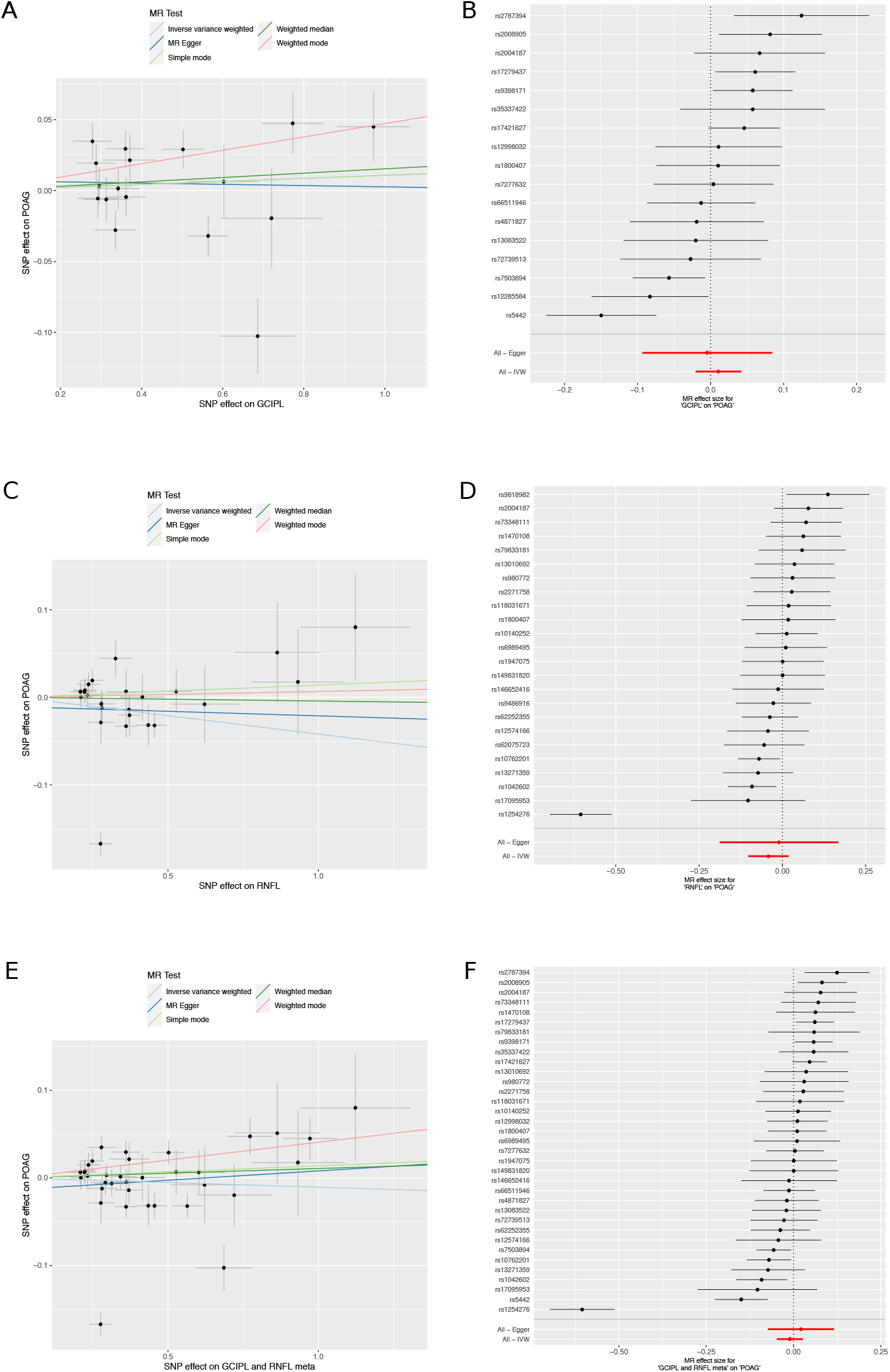
Mendelian Randomisation of Retinal Layers and Primary Open Angle Glaucoma. A) Scatter plot of the relationship between the effect size of SNPs found significantly associated with GCIPL thickness, and the effect size of those SNPs on POAG. B) Forest plot showing the effect size and direction of SNPs significantly associated with GCIPL thickness on POAG. C) Scatter plot of the relationship between the effect size of SNPs found significantly associated with RNFL thickness, and the effect of those SNPs on POAG. D) Forest plot showing the effect size and direction of SNPs significantly associated with RNFL thickness on POAG. E) Scatter plot of the relationship between the effect size of SNPs found significantly associated in the meta analysed GCIPL and RNFL thicknesses, and the effect of those SNPs on POAG. F) Forest plot showing the effect size and direction of SNPs significantly associated in the meta analysed GCIPL and RNFL thickness on POAG. POAG summary statistics were taken from the POAG International Glaucoma Genetics Consortium (IGGC) meta analysis [43].

**Figure 12.**
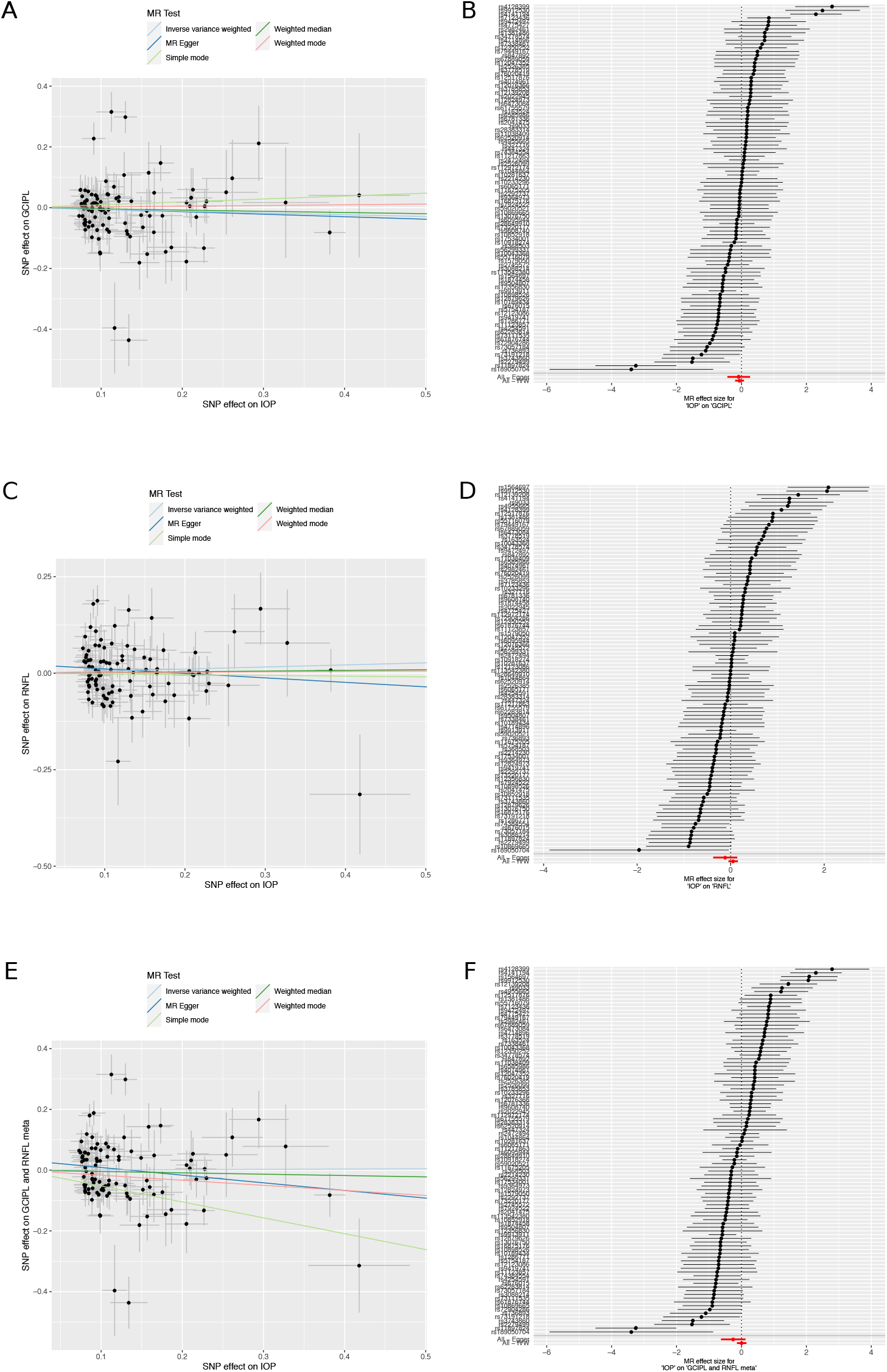
Mendelian Randomisation of Intraocualr Pressure and Retinal Layers. A) Scatter plot of the relationship between the effect size of SNPs found significantly associated with IOP, and the effect size of those SNPs on GCIPL thickness. B) Forest plot showing the effect size and direction of SNPs significantly associated with IOP on GCIPL thickness. C) Scatter plot of the relationship between the effect size of SNPs found significantly associated with IOP, and the effect of those SNPs on RNFL thickness. D) Forest plot showing the effect size and direction of SNPs significantly associated with IOP on RNFL thickness. E) Scatter plot of the relationship between the effect size of SNPs found significantly associated with IOP and the effect size of those SNPs in the meta analysed GCIPL and RNFL thickness. F) Forest plot showing the effect size and direction of SNPs significantly associated with IOP on the meta analysed GCIPL and RNFL thickness. Summary statistics for genetic association studies of IOP, were taken from [24].

**Figure 13.**
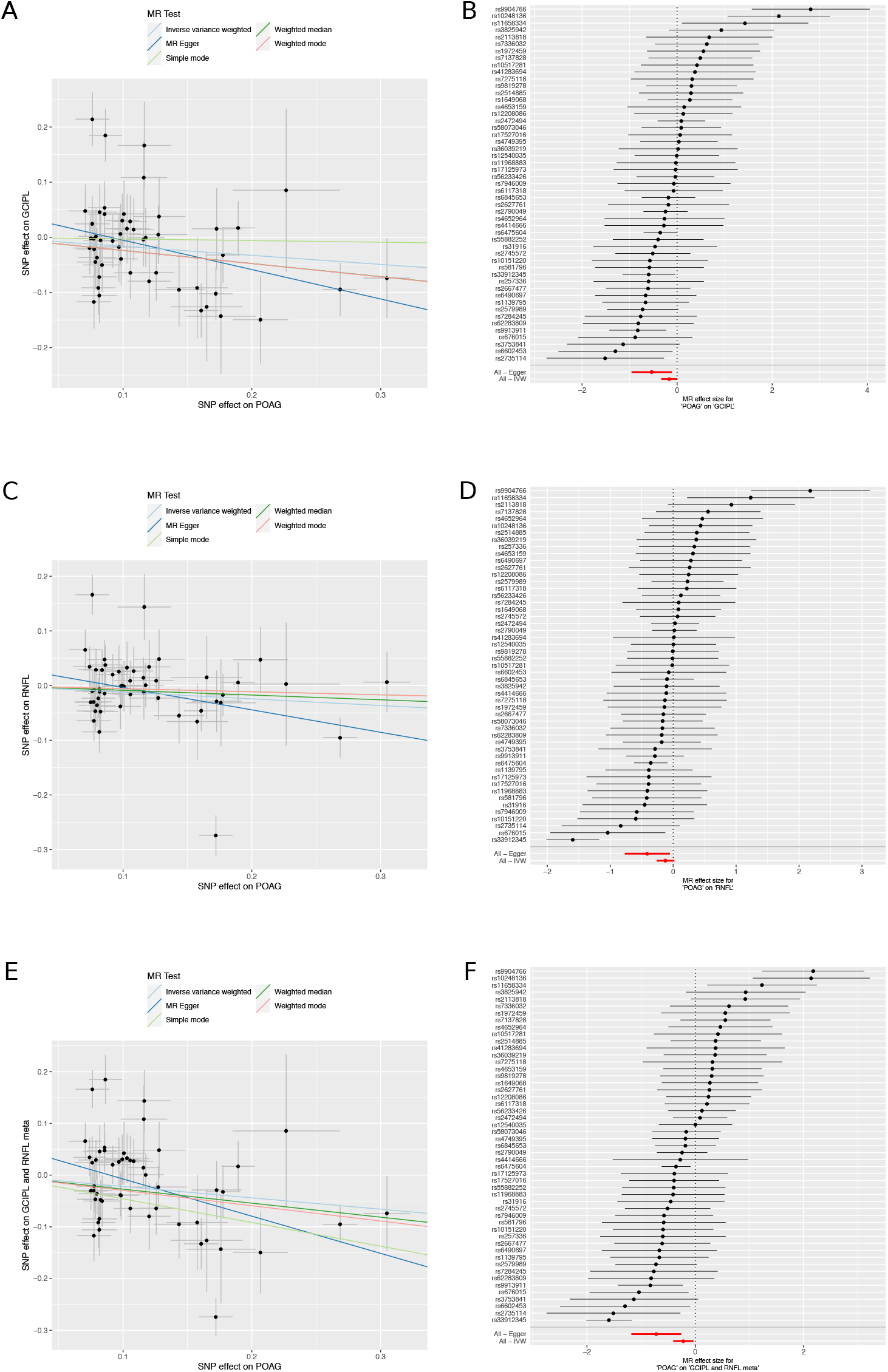
Mendelian Randomisation of Primary Open Angle Glaucoma and Retinal Layers. A) Scatter plot of the relationship between the effect size of SNPs found significantly associated with POAG, and the effect size of those SNPs on GCIPL thickness. B) Forest plot showing the effect size and direction of SNPs significantly associated with POAG on GCIPL thickness. C) Scatter plot of the relationship between the effect size of SNPs found significantly associated with POAG, and the effect of those SNPs on RNFL thickness. D) Forest plot showing the effect size and direction of SNPs significantly associated with POAG on RNFL thickness. E) Scatter plot of the relationship between the effect size of SNPs found significantly associated with POAG and the effect size of those SNPs in the meta analysed GCIPL and RNFL thickness. F) Forest plot showing the effect size and direction of SNPs significantly associated with POAG on the meta analysed GCIPL and RNFL thickness. POAG summary statistics were taken from the POAG International Glaucoma Genetics Consortium (IGGC) meta analysis [43].

The lack of concordance between RNFL or GCIPL genetically determined thickness and POAG is in contrast to their established use as diagnostic biomarkers for POAG [45, 46]. Consistent with previous epidemiological studies, the UK Biobank datasets shows thinner GCC for diagnosed glaucoma patients (Supplementary figure 7). The *SIX6* locus is the only locus clearly associated with GCIPL or RNFL and glaucoma. The same locus is also associated with a number of developmental traits, such as age of menarche and anthropometric traits. The role of the *SIX6* locus in development of the neural retina has been demonstrated in a zebrafish model, and a common missense variant Asn141His (rs33912345) has been implicated as causal [47].

## Discussion

We have performed the first large-scale genetic association study on inner retinal morphology. We explored a variety of options of how to model the phenotype, but with the large sample size present in UK Biobank, there was little difference in discovery power between a high dimensional perspective compared to the more straightforward GWAS of the measurements used in clinical practice, namely mean RNFL and GCIPL thickness. We robustly discovered 46 loci associated with at least one of the inner retinal thickness phenotypes across the genome. Many of the discovered loci are related to eye phenotypes; a notable association was to variants in genes associated with oculocutaneous albinism, which is known to affect the retinal pigment epithelium in the outer retina in addition to causing foveal hypoplasia [48]. Further exploration of these and one other locus shows that some minor foveal hypoplasia occurs in many individuals due to these variants. Although all three variants were associated with hair colour, the direction of effect on hair colour is not consistent with the direction of effect of foveal hypoplasia, showing that there is a complex relationship between the pigmentation pathway in retinal development compared to hair colour. The *TSPAN10* locus is less well described as being involved with pigmentation, with some evidence that it is involved in eye pigmentation specifically [49]. Ideally association to eye colour would also be tested, however the UK Biobank does not currently contain such phenotypic information. Although there is a complex genetic architecture for these three loci, it is clear that they collectively impact foveal development. Notably the three associated loci also showed significant association with visual acuity, implying a link between even subtle foveal hypoplasia and visual function; to our knowledge this is the first time genetic variation has been associated with visual acuity, as measured using a LogMAR chart. As the internal segmentation of the layers and the outline of the fovea were not available from the Topcon Advanced Boundary Segmentation (TABS) algorithm [50], the OCT segmentation software, it is complex to deconstruct which aspect of macular structure is changing the layer thickness measurements for these loci. The simplest explanation for these loci’s association is that the mild foveal hypoplasia caused by these variants is systematically changing the average GCIPL and RNFL measurements, potentially due to incorrect positioning of the macula within the Macula6 grid during scan acquisition. This suggests there is room for possible improvements in the image processing and measurement derivation. As these measurements are often used in clinical practice, in particular in the diagnosis of POAG, this retinal developmental variation will confound some uses of the current measurement schemes both in research and clinically.

The GWAS loci are also associated with a variety of other traits, with many loci also being associated with anthropometric traits, asthma, and blood cell related traits, and some neurological traits. This points to the broad and complex pleiotropy across biology but also suggests an opportunity for using the optically accessible retinal tissue, part of the central nervous system, as a source of potential biomarkers for other diseases [51, 52]. Consistent with this broad pleiotropy, GWAS loci are enriched in DNaseI hypersensitive sites in pancreas, kidney, blood and brain cells. The overlap with kidney tissue is interesting given the longstanding association between kidney and retinal disease [53, 54, 55]. This tissue overlap is also consistent with loci including rs2004187 (*IRX2*), which is associated with renal failure.

A surprise in our analysis was a consistent lack of overlap of our discovered loci with POAG loci, which has a large-scale consortium with a well-powered meta-analysis. The majority of our GCIPL and RNFL-associated variants did not associate with POAG in a large consortium meta-analysis. Furthermore, Mendelian randomisation tests did not support a causal effect of GCIPL or RNFL thickness on POAG. This suggests that the major genetic processes that underlie variation in inner retinal thickness do not affect POAG. As expected, a Mendelian randomisation test in the reverse direction did provide some weak evidence for the genetic processes underlying POAG causing inner retinal thinning. Also expected was the very striking Mendelian randomisation evidence for higher IOP causing POAG. Put together, these findings suggest that the genetic causes of POAG are primarily via raised IOP rather than processes underlying inner retinal thickness. As the thickness of these layers are consistently lower in individuals with POAG, this suggests that the change in thickness over time is a biomarker of the disease rather than the absolute level. This situation is similar to the observations that Hb1Ac is used as a biomarker for Type II diabetes but is confounded with red blood cell turnover and biology [56, 57]. In both this example and the case of inner retinal thickness, aspects of the biomarker biology influenced by genetics may confound the thresholds of the biomarkers for clinical use. Indeed, the discovered loci, particularly the three loci associated with foveal hypoplasia, are likely to be confounders of the POAG biomarker. In the future it may be possible to adjust for the developmental baseline of these layers using genetic markers to provide more accurate metrics for POAG incidence and progression. The lack of an unambiguous glaucoma definition within the UK Biobank, and the fact that the age of glaucoma onset is relatively late in comparison to the mean age of the UK Biobank dataset, currently limits this work in this cohort. In addition, access to longitudinal data, to allow for monitoring of changes in thickness with disease progression would allow for analysis to interrogate this theory further. Currently only very small datasets of such nature are available, but in the future this type of data may help further elucidate the pathways of well-established POAG genes.

This study focused on two measurements from the inner retina, widely used in clinical practice. We have characterised the genetics underlying these traits, illuminating their role in known eye diseases, and explored some of the developmental processes around the fovea. The images from which the phenotypes are extracted are available and likely can be processed to a far richer representation. Improvements in image analysis using deep learning techniques [58] show great promise in this regard. We will extend this work both to the outer retina and to these richer phenotypes, increasing the breadth and detail of eye morphology that can be explained.

## Methods

### UK Biobank cohort

The UK Biobank is a large, well-studied prospective cohort sampled from across the UK at various sites. Participants completed a baseline questionnaire, physical measurements and provision of biological samples. The questionnaire collected information on demographics, anthropometrics, socioeconomic and lifestyle factors. This study was conducted between 2006 and 2010, recruiting more than half a million people aged 40-69 years old, identified via the NHS registry [59]. The study was conducted with the approval of the North-West Research Ethics Committee (ref 06/MRE08/65), in accordance with the principles of the Declaration of Helsinki, and all participants gave written informed consent. This research has been conducted using the UK Biobank Resource under Application Number 2112.

### Ophthalmic measurements and Optical Coherence Tomography

A subset of the cohort, comprising 132,041 participants, had further ocular data collected including IOP, visual acuity and autorefraction. As part of this ophthalmic assessment, 67,321 individuals underwent macular spectral domain OCT (SD-OCT) imaging.

### OCT Imaging

The SD-OCT imaging was done using the Topcon 3D OCT1000 Mark II and was performed following visual acuity, autorefraction and IOP measurements. The SD-OCT imaging was carried out in a dark room without pupil dilation using the 3-dimensional 6×6 mm^2^ macular volume scan mode (512 A scans per B scan; 128 horizontal B scans in a raster pattern). The right eye was imaged first, and the scan was repeated for the left eye in most individuals.

### Visual acuity measurement

Visual acuity was measured in both eyes using a logarithm of the minimum angle of resolution (LogMAR) chart (Precision Vision, LaSalle, Illinois, USA) displayed on a computer screen. The test was carried out with participants wearing their distance glasses at 4 m, or at 1 m if a participant was unable to read letters at 4 m. Participants were asked to read from the top of the chart and the test was terminated when 2 or more letters were read incorrectly. For patient-level visual acuity, we considered the value for the eye with better visual acuity.

### Autorefraction measurement

Non-cycloplegic autorefraction was carried out using the Tomey RC-5000 Auto Refractor Keratometer (Tomey Corp., Nagoya, Japan).2 Up to 10 measurements were taken for each eye and the most reliable measure was automatically recorded. Spherical equivalent was calculated as spherical power plus half cylindrical power.

### Derivation of retinal thickness measures

The OCT images were stored as both .fds files, a proprietary image storage file format, as well as as .dicom files. Version 1.6.1.1 of the Topcon Advanced Boundary Segmentation (TABS) algorithm [50] was applied to the images to segment the various retinal layers, and to calculate the thickness of such layers across the retinal fields, or across sub-fields as defined by the Early Treatment of Diabetic Retinopathy Study (ETDRS) [60] or Macula6 grid [61](Figure 1D).

### Quality control and inclusion/exclusion criteria

Participants were excluded from the study if they had withdrawn their consent, or were recommended for exclusion from genetics studies by UK Biobank (Figure 1C). Participants that were included represent the densest populated well-mixed population within the overall dataset. These participants were identified as being within a defined euclidean distance of the mean of the desired population, as selected by comparison to the HapMap Phase III study [62], in the PC1-PC2 space. Individuals were then excluded if they were related to third degree or more, based on the kinship information provided as a UK Biobank variable [63]. Further participants were removed from the dataset based on rigorous quality control of their OCT scans using previously implemented methods [28]. Briefly, this entailed all OCT images with an image quality score less than 45 being removed from the data set. A number of other segmentation indicators were also used as quality control metrics and for each of these, individuals representing the poorest 20% of the population in each of these measures were removed from the dataset. These segmentation indicators included: Inner Limiting Membrane (ILM) Indicator, a measure of the minimum localised edge strength around the ILM boundary across the entire scan. ILM indicator is useful for identifying blinks, scans that contain regions of severe signal fading, and segmentation errors; Valid count, a measure used to identify scans with a significant degree of clipping in the OCT scan’s z-axis dimension; Minimum motion correlation, maximum motion delta and maximum motion factor, these indicators use both the nerve fibre layer and the full retinal thicknesses, from which Pearson correlations and absolute differences between the thickness data from each set of consecutive B-scans are calculated. The lowest correlation and the highest absolute difference in a scan are the resulting indicator scores and serve to identify blinks, eye motion artefacts, and segmentation failures. It should be noted that the various indicators, including the image quality score, tend to be highly correlated with one another. The participants were further filtered to remove participants with outlier refractive error values defined as data points lying outside one standard deviation of 1.5 times the inter-quartile range. The final dataset consisted of 31,434 participants.

### Genome-wide Association Study

GWAS were performed using an additive linear model implemented using BGENIE v1.3 [64]. An individual GWAS was conducted for each of the two phenotypes, representing the thickness of the RNFL and GCIPL respectively, both averaged across the Macula6 grid. Eye-specific covariates, namely refractive error, calculated from the spherical and cylindrical volume (refractive error = spherical power + 0.5 × cylindrical power), and technical covariates (Macula centre frame, Macula centre aline, ILM indicator, Valid count, Minimum motion correlation, Maximum Motion Delta, Maximum motion factor and Image quality) were regressed out of the thickness measurements for the separate eyes before the phenotypic measure of the mean across left and right eyes was calculated. Age, weight, height, sex, the ID of the OCT machine used in scan acquisition and the first 20 genotype PCs were then used as covariates in the model. SNPs were considered significantly associated if they met the consensus genome-wide significance level (P <5 × 10^−8^). LD-score regression was implemented using LD SCore v1.0.1 [65].

### Discovered Associated SNP Set Creation

MTAG [31] was used to perform multi trait meta analysis across the GWAS summary statistics from the RNFL and GCIPL thickness analyses. This produced new adjusted P-values for each trait. For each SNP, the lowest P-value across the two traits was selected as the meta P-value. This was then used to plot a meta manhattan plot (Figure 2). GCTA Conditional and Joint Analysis (COJO) [66] was used to perform step-wise model selection to select significantly associated independent loci that were more than 10Mb apart. The resulting 46 loci constituted our discovered associated SNP set.

### Annotation of variants

Each SNP in the discovered associated SNP set was manually annotated using both ENSEMBL [67] and the Open Targets Genetics [68] PHeWAS annotations. SNPs were further filtered and labelled as being within the same loci if they were within 1.5Mb of one another. This is represented in the shading of loci within Table 1, with loci considered to be within the same loci shaded the same colour, alternating grey and white.

### Replication of inner retinal morphology GWAS and examination of association with POAG

Replication of the primary GWAS was sought in two datasets, the Raine Study, and the Rotterdam study.

The Raine Study is a multigenerational, longitudinal study based in Perth, Western Australia [69]. At the 20-year follow-up of the cohort, 1344 participants were enrolled in Raine Study Gen2, a cross-sectional study of eye diseases in young adults [70]. All participants underwent a standardised ocular examination, which included optical coherence tomography (OCT; Heidelberg Spectralis, Heidelberg Engineering GmbH, Heidelberg, Germany) measurements of the macula using a raster scan (31-line horizontal scan, 30° × 25°) centred on the fovea. Automatic segmentation was performed with the Spectralis software. Right and left eyes data were averaged for each subject. Genotyping was performed on 1593 participants, which included those who did not attend the vision assessments, via the Illumina Human 660W-quad BeadChip (Illumina, Inc., San Diego, CA, USA). After standard quality control, the cleaned genotypic datasets included 1495 individuals. Details of the quality control step have been previously described [71]. The mean macula RNFL (mRNFL) value was obtained by averaging the RNFL value on both eyes. We derived the GCIPL phenotype by summing up mean values of GCL and IPL. After removing individuals with missing phenotypes, the GWAS analyses on mRNFL (n=1014) and GCIPL (n=1025) for the Raine cohort was conducted using Plink 2.00alpha. We adjusted the mean mRNFL values by the participant’s average axial length (both eyes), fitted along with other standard covariates including age, sex and the top 5 ancestral principal components. The summary statistics for the relevant requested SNPs for replication were supplied back to the consortium.

The Rotterdam Study (RS) is a prospective population-based cohort study among individuals 45 years or older, residing in Ommoord, a district in Rotterdam, the Netherlands [72]. The first cohort started in January 1990 (RS,1 n = 7983). In February 2000 (RS2, n = 3011) a second and in February 2006 (RS3, n = 3932) a third cohort was started. Follow-up examinations took place every 3 to 4 years. In September 2007, spectral-domain OCT scanning was added to the protocol. The current analysis comprises OCT data acquired at the fifth visit of the first cohort (RS1), the third visit of the second cohort (RS2), and the second visit of the third cohort (RS3). Eyes were initially scanned with the spectral-domain OCT-1000 Mark 2 (Topcon, Tokio, Japan). From August 2011 onwards, this device was replaced with the spectral-domain OCT-2000 owing to an update. As a result of this update about half of the RS2 cohort and all persons of RS3 cohort were examined on the OCT-2000 machine. The macula was scanned in the horizontal direction in an area of 6 × 6 × 1.68 mm with 512 × 512 × 480 voxels (using OCT-1000) and 6 × 6 × 2.30 mm with 512 × 512 × 885 voxels (using OCT-2000). Macula volumes scans were segmented using Iowa Reference Algorithms, version 3.6 (Retinal Image Analysis Lab, Iowa Institute for Biomedical Imaging, Iowa City, IA) (available at https://www.iibi.uiowa.edu/content/shared-software-download) [73]. Indices of quality control were used to preserve good-quality images and to exclude scans with segmentation errors. Scans included in our study had a segmentability index >20% and an undefined region of <20% (measures of segmentation/algorithm failures). If the scan of both eyes passed quality and control a random scan was chosen for the analysis. A total of 1000 persons in RS1, 1448 persons in RS2 and 765 persons in RS3 passed quality and control and were included in the study. The volume of the ganglion cell complex (RNFL + GCL + IPL) was calculated for 6×6mm macular surface and used as a quantitative phenotype in the GWAS. Genotyping was performed on either Illumina 550 (+duo) or Illumina 610 quad (Illumina, Inc., San Diego, CA, USA), genotypes were imputed to haplotype reference consortium (HRC) release 1 imputation panel, a detailed description of genotyping and pre- and post-imputation quality and control have been described elsewhere [74, 75]. For the GWAS, genotype and phenotype data were available for 899 persons in RS1, 1131 persons in RS2 and, 326 persons in RS3. We conducted a GWAS in PLINK v2.00 alpha and fitted a linear model adjusted for spherical equivalent, sex, age and the top 5 ancestral principal components [76]. As two different types of OCT machines were used during the data collection we initially analysed the cohorts separately and stratified by OCT machine. The results of these genome-wide analyses were then combined in a meta-analysis using METAL [77]. For every SNP we assessed the level of heterogeneity by calculating I2 values and Cochrans Q-test for heterogeneity as implemented in METAL.

### Higher dimensional detailed retinal phenotyping

The surface of the inner limiting membrane (ILM) and bruchs membrane (BM) were segmented for each scan. To do so vectors of the columns, or axial scans, of an image slice were smoothed using a running median. Points of interest were identified as those outside two standard deviations of the median. The maximum and minimum of these points of interest were labelled as the ILM and BM, respectively. This method was iterated across each axial scan and outlier interpolation was applied to the composite layer boundary.

The coordinates of the foveola in the *x, z* dimension were calculated by finding the minima of *y* across each of these dimensions. The mean intensity value for each *x* position in a line scan was calculated for every line scan, producing a matrix where *y* is the mean intensity value within each line scan and *x* is the line scan index. This matrix was collapsed by taking a mean across line scan indices. The local minima, confined by two local maxima, was labelled as the *z* coordinate of the foveola. The same process was repeated in the perpendicular axes to identify the *x*-coordinate of the foveola. The coordinates of the foveola were used to centre the images within set dimensions (650 × 512 × 128), so that the foveola was in the centre and the overhanging edges were cropped. For each image the matrix was calculated representing the distance between the ILM surface and the BM.

### Association with foveal hypoplasia measure and pigmentation

For each genetic variant, the population was stratified by dosage of the alternative allele (0, 1 and 2). Means of the retinal thickness matrices, considering left and right eyes separately, were made across the participants within each genotype group resulting in the mean matrix per genotype per eye. A matrix of the difference in retinal thickness between homozygous reference and heterozygous, and homozygous reference and homozygous alternative, were calculated. Heatmaps of these difference matrices were plotted, with the colour representing the thickness of the retina. Additionally, a spline function is applied to the cross sectional vector along both axes of the models for each genotype and plotted as a line graph.

The overall thickness at the central point for the fovea was also calculated for the left eye of each individual. This was defined as the overall thickness of the retina at the midpoint of the two cross-sectional vectors, one across A-scans, one across B-scans. These values were used as input to a linear model looking at the effect of each of the variants in the discovery set on the overall retinal thickness at the fovea. SNPs were considered to have a significant effect on the thickness of overall retinal thickness if P <0.05 following correction for Bonferroni correction for multiple testing for both retinal thickness values. The significant subset of SNPs were used in linear models of the SNP on hair and skin colour, as self-reported in the UK Biobank. In the case of hair colour, those with red hair were removed from the analysis. Both hair colour and skin colour were coded numerically, as in the UK Biobank, in ascending order from light to dark. SNPs were considered to have a significant effect on hair colour or skin colour if P <0.05 after correction for multiple testing.

### Association with visual acuity

The lead inner retina-association variants were tested for association with visual acuity. LogMAR visual acuity in the better seeing eye was used as the outcome measure, and the linear model run across all Europeans, as defined by genetic PCs (n=100,818), and adjusted for age, sex, and the first 15 genetic principal components.

### Mendelian Randomisation Analysis

Mendelian randomisation analysis was undertaken using the TwoSampleMR package in R [78]. In the MR analysis, SNPs from the discovered associated SNP set were used as the exposure variable. Summary statistics for genetic association studies of IOP, were taken from [24] and when used as an exposure variable, were selected for genome-wide significance (P <5 × 10^−8^). The internal LD pruning function was also applied to all exposure variables. POAG summary statistics were used from the POAG International Glaucoma Genetics Consortium (IGGC) meta analysis [43]. The GCIPL and RNFL thickness (whether used as an exposure or an outcome) had the same set of technical covariates applied as the GWAS listed above. Eye-specific covariates, refractive error, calculated from the spherical and cylindrical volume (refractive error = spherical power + 0.5 × cylindrical power), and technical covariates (Macula centre frame, Macula centre aline, ILM indicator, Valid count, Minimum motion correlation, Maximum Motion Delta, Maximum motion factor and Image quality), were regressed out of the thickness measurements for the separate eyes before the phenotypic measure of the mean across left and right eyes was calculated. The association was applied with age, weight, height, sex, the ID of the OCT machine used in scan acquisition and the first 20 genotype PCs as covariates in the linear model. Although some of the associated SNP instruments were positive for these covariates (in particular height and weight) we considered these to be likely examples of horizontal pleiotropy. We used two meta-analysis methods, MR-Egger and inverse variance weighting (IVW); as the samples for the IOP consortium and the POAG consortium are independent of UK BioBank, the assumptions made by the MR-Egger scheme are valid.

### GCC thickness in the UK Biobank glaucoma population

The cohort used in the GWAS was divided into those that had glaucoma (n=2751), and those that did not (n=28,314). Glaucoma case status was ascertained as participants who reported a history of glaucoma, glaucoma laser or glaucoma surgery on a touchscreen questionnaire or participants who had a glaucoma-related ICD 9/10 code on linked hospital episode statistic data (ICD 9: 365.*; ICD 10: H40.* [excluding H40.0], H42.*). GCC thickness, defined as the sum of GCIPL and RNFL thickness was calculated and a mean taken across the two eyes. A Wilcoxon rank sum test was performed to test for significant difference in GCC thickness between the two groups.

## Data Availability

The genetic and phenotypic UK Biobank data are available upon application to the UK Biobank.

## Data availability

The genetic and phenotypic UK Biobank data are available upon application to the UK Biobank.

## Acknowledgements

HC and EB are funded by EMBL. APK is supported by a Moorfields Eye Charity Career Development Fellowship. The Rotterdam Study is funded by Erasmus MC and Erasmus University, Rotterdam, Netherlands Organization for the Health Research and Development (ZonMw), the Research Institute for Diseases in the Elderly (RIDE), the Ministry of Education, Culture and Science, the Ministry for Health, Welfare and Sports, the European Commission (DG XII), and the Municipality of Rotterdam. The authors are extremely grateful for the selfless participation of individuals in all cohorts used in this study (the UK BioBank, The Rotterdam Study and the Raine Study), and the staff managing these cohorts. LP is funded by NIH EY015473. JW is funded by NIH/NEI P30 EY014104 and R01 EY022305. AH is supported by an Australian National Health and Medical Research Council Practitioner Fellowship. EJ and HC are supported by National Eye Institute (NEI) grant R01 EY027004 and by the National Institute of Diabetes and Digestive and Kidney Diseases (NIDDK) R01 DK116738.

## Author Contribution

HC conceived the analysis, performed the analysis and wrote the paper. PH and TF provided advice on the analysis and wrote the paper. DM, JO, JC, DA, LRP, AWH, PWMB, AAHJT, CCWK, and SY provided replication. APK provided analysis on visual acuity. PJF, CAR and PJP coordinated the consortium work and helped in segmentation of OCT images. EB and APK conceived the analysis and oversaw the work. All authors reviewed the final manuscript.

## Supplementary Figures

## Supplementary Tables

